# Beyond the Spectrum: Subtype-Specific Molecular Insights into Autism Spectrum Disorder Via Multimodal Data Integration

**DOI:** 10.1101/2024.09.17.24313857

**Authors:** Javad Zahiri, Mehdi Mirzaie, Kuaikuai Duan, Yaqiong Xiao, Caitlin Aamodt, Xiaotong Yang, Sanaz Nazari, Charlene Andreason, Linda Lopez, Cynthia Carter Barnes, Steven Arias, Srinivasa Nalabolu, Lana Garmire, Tianyun Wang, Kendra Hoekzema, Evan E. Eichler, Karen Pierce, Nathan E. Lewis, Eric Courchesne

**Author notes:** Correspondence to: Javad Zahiri, Autism Center of Excellence, Department of Neurosciences, University of California, San Diego, La Jolla, CA, USA, Nathan Lewis, Department of Pediatrics, University of California, San Diego, La Jolla, CA, USA, Eric Courchesne, Autism Center of Excellence, Department of Neurosciences, University of California, San Diego, La Jolla, CA, USA. Co-Senior Author (Karen Pierce, Nathan E. Lewis). Senior Author (Eric Courchesne).

## Abstract

Some toddlers with autism spectrum disorder (ASD) have mild social symptoms and developmental improvement in skills, but for others, symptoms and abilities are moderately or even severely affected. Those with profound autism have the most severe social, language, and cognitive symptoms and are at the greatest risk of having a poor developmental outcome. The little that is known about the underlying biology of this important profound autism subtype, points clearly to embryonic dysregulation of proliferation, differentiation and neurogenesis. Because it is essential to gain foundational knowledge of the molecular biology associated with profound, moderate, and mild autism clinical subtypes, we used well-validated, data-driven patient subtyping methods to integrate clinical and molecular data at 1 to 3 years of age in a cohort of 363 ASD and controls representative of the general pediatric population in San Diego County. Clinical data were diagnostic, language, cognitive and adaptive ability scores. Molecular measures were 50 MSigDB Hallmark gene pathway activity scores derived from RNAseq gene expression. Subtyping identified four ASD, typical and mixed diagnostic clusters. 93% of subjects in one cluster were profound autism and 93% in a different cluster were control toddlers; a third cluster was 76% moderate ability ASD; and the last cluster was a mix of mild ASD and control toddlers. Among the four clusters, the profound autism subtype had the most severe social symptoms, language, cognitive, adaptive, social attention eye tracking, social fMRI activation, and age-related decline in abilities, while mild autism toddlers mixed within typical and delayed clusters had mild social symptoms, and neurotypical language, cognitive and adaptive scores that improved with age compared with profound and moderate autism toddlers in other clusters. In profound autism, 7 subtype-*specific* dysregulated gene pathways were found; they control embryonic proliferation, differentiation, neurogenesis, and DNA repair. To find subtype-*common* dysregulated pathways, we compared all ASD vs TD and found 17 ASD subtype-*common* dysregulated pathways. These *common* pathways showed a severity gradient with the greatest dysregulation in profound and least in mild. Collectively, results raise the new hypothesis that the continuum of ASD heterogeneity is moderated by subtype-*common* pathways and the distinctive nature of profound autism is driven by the differentially added profound subtype-*specific embryonic pathways*.

## 1. Introduction

ASD is 80% to 90% heritable, and yet the molecular pathobiology underlying its social symptoms at the individual patient level at early ages remains speculative and largely a mystery. Despite the high heritability (Bai *et al*., 2019), the genetic basis is unknown for an estimated 90% of ASD patients (Feliciano *et al*., 2019). Gene diagnostic panels have poor clinical utility giving diagnostic yields ranging from a low of 0.22% to a high of only 10% (Ní Ghrálaigh *et al*., 2023a). De novo variants (DNV) explain only ∼2% of variance in ASD (Gaugler *et al*., 2014; Satterstrom *et al*., 2020), and a recent study concluded “a continued focus on DNVs for ASD gene discovery may yield diminishing returns” (Zhou *et al*., 2022). Even for the small 10-13% subset of ASD patients who have a mutation in a risk gene, how or even whether a mutation causes the social symptoms is unknown for hundreds of hypothesized “risk” mutations, and no single ASD child with one of those mutations represents even a tiny fraction of all the candidate risk mutations reported. Thus, since such DNA mutations are so rare, none are representative of the ASD population, and the ASD molecular landscape computationally built from rare single cases does not represent the ASD population. Therefore, despite hundreds of millions of research dollars and hundreds of papers, still missing is not only knowledge of the molecular pathobiology of ASD social symptoms, but also of the molecular pathobiology underlying the known diversity in ASD symptom presentation, progression and outcome at early ages. It is at early ages when such knowledge could have the greatest benefit for affected toddlers. As such, the ASD DNA mutation field is largely empty of critical information about ASD clinical-molecular subtypes and is at a standstill.

A new in vivo (blood) gene expression study identified a pathobiological signature that accurately diagnoses ASD individuals from among typical toddlers by combining large numbers of dysregulated gene expression patterns; AUC-ROC and AUC-PR range from 84% to 92% (Bao *et al*., 2023), which is vastly superior to gene mutation panels (Ní Ghrálaigh *et al*., 2023a). The transcriptomic diagnostic signature included gene expression dysregulations that enriched cell cycle mechanisms and gene signaling pathways (PI3K-AKT, RAS-ERK and Wnt) that regulate multiple prenatal neurodevelopmental processes (Courchesne *et al*., 2019; Gazestani *et al*., 2019). Importantly, the study also examined whether the presence of ASD “risk” gene mutations could improve diagnostic accuracy. Contrary to expectations, DNA mutations in SFARI Level 1 and 2 ASD risk genes were as common in typical (11%) as in ASD toddlers (11%), suggesting those genes may not be risk-relevant and can lead to chance diagnostic classifications, as predicted from the Ni Ghralaigh *et al*. study (Ní Ghrálaigh *et al*., 2023b). A different study found that a dysregulated gene network composed of those three signaling pathways was significantly overactive in vivo in ASD individuals relative to typicals (Gazestani *et al*., 2019). The degree of overactive expression of this network was related to social symptoms severity in ASD. To our knowledge, no risk gene mutation finding has shown such relationships to core ASD social symptoms in large ASD samples at early ages. That same study showed this social symptom-relevant network was also significantly overactive in ASD prenatal neurons and neural progenitor cells derived from ASD toddlers with early brain overgrowth that is correlated with excess cell proliferation; this multi-pathway network is upregulated in the first and second trimesters during proliferation and neurogenesis (Gazestani *et al*., 2019; Courchesne, Gazestani and Lewis, 2020).

A large literature on computational systems biology, cell and animal model, postmortem and GWAS research also implicates prenatal dysregulation of cell proliferation, neurogenesis, and corticogenesis as well as dysregulation in these three signaling pathways (Courchesne *et al*., 2019; Courchesne, Gazestani and Lewis, 2020). However, this larger literature has not linked such multi-pathway, multi-process biological evidence with clinical symptoms in living ASD patients. Studies and reviews of the ASD blood gene expression literature also found dysregulated gene expression in a several pathways and processes, including PI3K-AKT-mTOR, RAS signaling pathways, ribosomal translation signal, cell cycle, neurogenesis, gastrointestinal disease, immune/inflammation, interferon signaling, and the natural killer cytotoxicity pathway (Gregg *et al*., 2008; Enstrom *et al*., 2009; Kong *et al*., 2012; Ch’ng *et al*., 2015; Pramparo, Lombardo, *et al*., 2015; Pramparo, Pierce, *et al*., 2015; Diaz-Beltran, Esteban and Wall, 2016; Ansel *et al*., 2017; Tylee *et al*., 2017; He *et al*., 2019; Lee *et al*., 2019), but again such molecular evidence has seldom been linked with ASD clinical characteristics, developmental change, and subtypes.

Lastly, ASD children with the most severe social and language symptoms –profound autism^5,6^– are at the greatest risk for a poor lifelong outcome. Due to low levels of participation in research studies and challenges in early-age identification, little is known about the underlying molecular biology in this important group of autistic toddlers. Because severe impairments in social perception and reactivity and language are early and specific signs of profound autism, there is a particular necessity for non-invasive approaches to uncover the molecular biology associated with profound autism and how it may differ from moderate and milder autism. Insight into embryogenic dysregulation of brain growth that distinguishes profound ASD from mild ASD clinical phenotypes, comes from a new brain cortical organoid (BCO) study^4^. That study shows that as early as embryogenesis, the biological bases of two subtypes of ASD social and brain development –profound autism and mild autism– are already present and measurable and involve dysregulated cell proliferation and accelerated neurogenesis and growth. The larger the embryonic BCO size in ASD, the more severe the toddler’s social symptoms and the more reduced the social attention, language ability, and IQ, and the more atypical the growth of social and language brain regions. This suggests the molecular dysregulations underlying profound autism differ in type or degree and might be detectable at very early postnatal ages when we find profound autism social brain activity and clinical features are robustly identified as a distinct subtype from other ASD toddlers (Xiao *et al*., 2022; Taluja *et al*., 2024).

Here we identify molecular pathobiology subtypes underlying ASD patient subtypes of early age clinical presentation, progression and clinical outcome. To do so, we use an established precision medicine method, Similarity Network Fusion (SNF), that integrates different modalities as single Patient Similarity Network (PSN). Using this PSN, we can identify clusters of patients and controls whose social and language clinical and molecular features are maximally similar to each other and maximally different from those in other clusters. This increases power to detect subtype-relevant molecular pathobiology associated with profound autism and milder clinical phenotypes. To objectively pinpoint and quantify dysregulated molecular pathways in each identified ASD patient subtype, we use the Hallmark pathways from MSigDB^5^. Gene sets in Hallmark pathways utilize a resource of tens of thousands of gene sets and are linked with gene expression data refining and validating Hallmark pathway signatures. Additionally, we evaluate differential expression of both small and long non-coding RNAs within each subtype.

## 2. Method

### 2.1 Participant recruitment

We performed ASD subtyping using similarity network fusion based on 12 different clinical and transcriptomic features of 363 male toddlers aged 1–4 years. The Institutional Review Board approved this study at the University of California, San Diego. According to the Declaration of Helsinki, parents provided written informed consent and were paid for their participation. Identically to the approach used in our other recent studies(Gazestani *et al*., 2019; Xiao *et al*., 2022; Bao *et al*., 2023), toddlers were recruited through two mechanisms: community referrals (website, social media, etc.) or a general population-based screening method called the 1-Year Well-Baby Check-Up Approach (now called the Get S.E.T. Early model)^7^ that allowed the prospective study of ASD beginning as young as ages 12 to 24 months, on the basis of a toddler’s failure of the Communication and Symbolic Behavior Scales Developmental Profile (CSBS-DP) Infant-Toddler Checklist. All toddlers, including control subjects, received a battery of standardized psychometric tests by highly experienced Ph.D. level psychologists including the Autism Diagnostic Observation Schedule score (ADOS) (Module T, 1 or 2), the Mullen Scales of Early Learning, and the Vineland Adaptive Behavior Scales. Testing sessions routinely lasted 4 hours and occurred across 2 separate days. Toddlers younger than 36 months at the time of initial clinical evaluation were followed longitudinally approximately every 9 months until a final diagnosis was determined at age 2–4 years. Toddlers were categorized into two groups based on their final diagnosis assessment: ASD (N=152), children with the diagnosis of ASD, and nonASD toddlers (N=211). The nonASD group includes typically developing (TD) (N=137), delayed toddlers (DL) including language delay, global developmental delay, motor delay and non-autistic ASD-feature toddlers (N=74). Table 1 shows the detailed clinical characteristics of these toddlers.

**Table 1:**
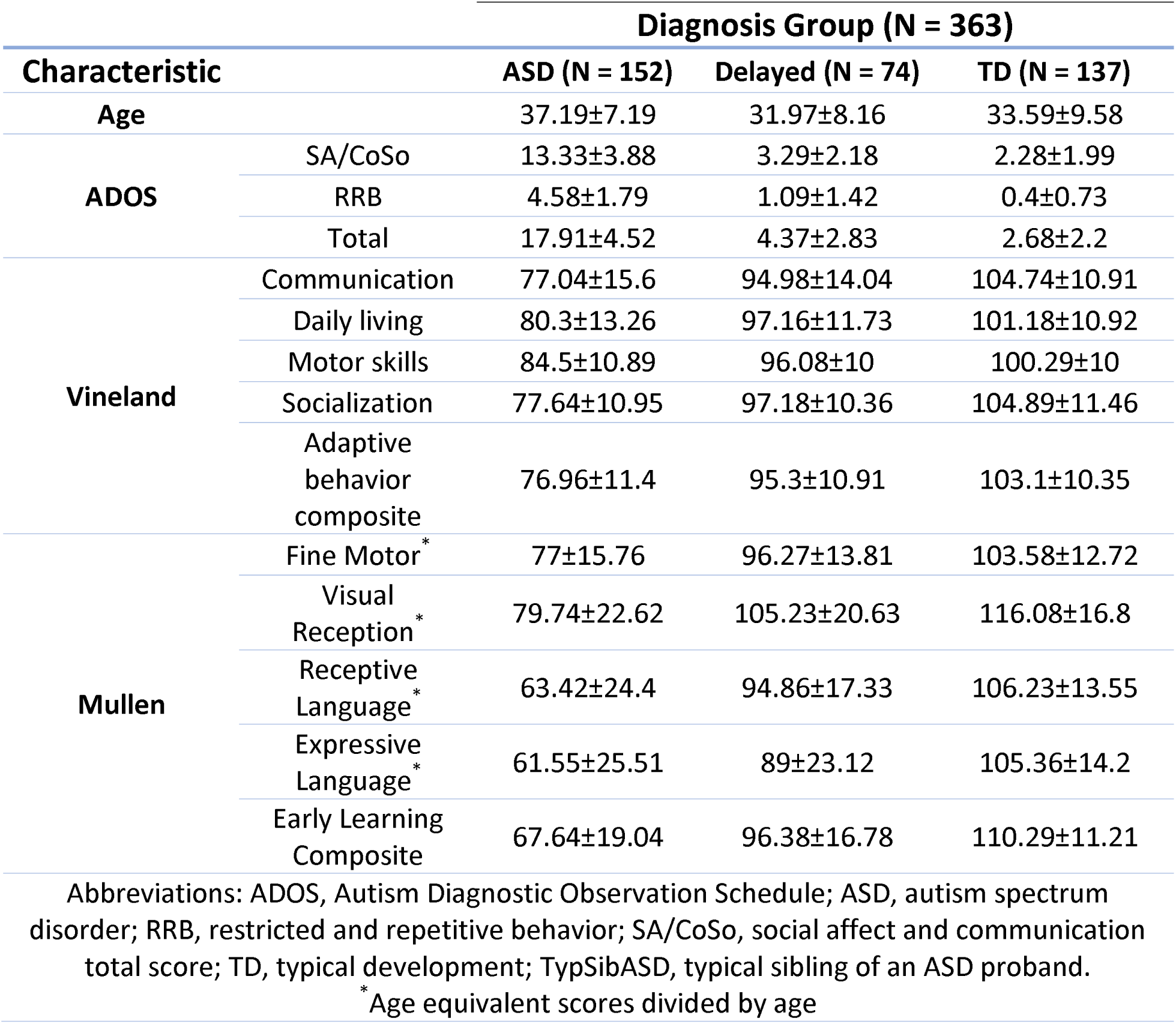
Clinical Characteristics of Toddlers Used in This Study.

### 2.2 Blood sample collection and RNA extraction

Four to six milliliters of blood were collected into EDTA-coated tubes from toddlers on visits. The blood draw was done only if toddlers had no fever, cold, flu, infections, or other illnesses, or use of medications for illnesses 72 h before the blood draw. Blood samples were passed over a LeukoLOCK filter (Ambion) to capture and stabilize leukocytes and immediately placed in a –20 °C freezer. Total RNA was extracted according to standard procedures and manufacturer’s instructions (Ambion). LeukoLOCK disks (Ambion cat no. 1933) were freed from RNAlater, and Tri-reagent (Ambion cat no. 9738) was used to flush out the captured lymphocytes and lyse the cells. RNA was subsequently precipitated with ethanol and purified through washing and cartridge-based steps. The quality of the mRNA samples was quantified according to the RNA Integrity Number (RIN): values of 7.0 or greater were considered acceptable, and all processed RNA samples passed RIN quality control. Quantification of RNA was performed with a NanoDrop spectrophotometer (Thermo Scientific). Samples were prepared in 96-well plates at a concentration of 25 ng/μl.

### 2.3 RNA Sequencing Plate design

Large RNA sample studies are vulnerable to batch effects across plates that can eliminate biological expression effects. To minimize batch effects, we used our BalanceIT tool (Chiang *et al*., 2021) in the plate design procedure. BalanceIT uses a genetic algorithm to organize samples into batches of plates before sequencing so that all are optimally balanced for 20 potentially confounding factors (e.g., age, gender, race, ethnicity, diagnosis, IQ, etc.) within and across plates.

#### Sequencing

The extracted bulk RNA has been sequenced using a NovaSeq 6000 platform in the Institute for Genomic Medicine (IGM) Center at the University of California, San Diego. The final RNA-seq reads were paired-end 151-base-pair length.

### 2.4 RNA-Seq preprocessing, mapping, and expression quantification

The read quality was checked with FastQC and low-quality bases and adapters were removed using trimmomatic^8^. Then, reads were aligned to the grch38 human reference genome using HISAT2^9^. HISAT2 mapping results were sorted according to genomic locations using samtools^10^, and genetic feature abundance was quantified using StringTie (Pertea *et al*., 2015). Transcriptome assembly was done using the merge function from StringTie tool. Then prepDE function was used to extract read counts directly from the genetic feature format (gtf) files generated by StringTie. Subsequently, ComBat-seq (Zhang, Parmigiani and Johnson, 2020) was used for batch effect adjustment. Batch corrected counts were normalized according to the library size using edgeR. Prior to doing differential expression analysis, low expressed genetic features were removed. Finally, we used the generalized linear model implemented in edgeR to find differentially expressed (DE) genes. We applied this pipeline for quantifying the expression values of three groups of genes: protein-coding, long non-coding RNA (lncRNA), and small non-coding RNA (sncRNA) including micro-RNA, scaRNA, snoRNA, and snRNA.

### 2.5 Pathway activity scores

We employed gene set variation analysis (GSVA) (Hänzelmann, Castelo and Guinney, 2013) to assess the activity of the 50 hallmark pathways defined by Broad institute and UCSD in the Molecular Signatures Database (MSigDB) (Liberzon *et al*., 2015). To calculate gene set activity scores for each pathway in each subject based on the normalized gene expression values, we utilized the ‘gsva’ R package. We used this package under the default settings except for the kcdf parameter. This parameter was set to “Poisson” as it is better suited for handling RNA-seq read counts.

### 2.6 Similarity network fusion and subtyping

#### 2.6.1 Patient Similarity Networks

Twelve different patient similarity networks (PSNs) have been constructed using eight different clinical measures, which we have denoted as “iSNF” clinical measures, and four transcriptomic PSNs (Figure 1).

**Figure 1:**
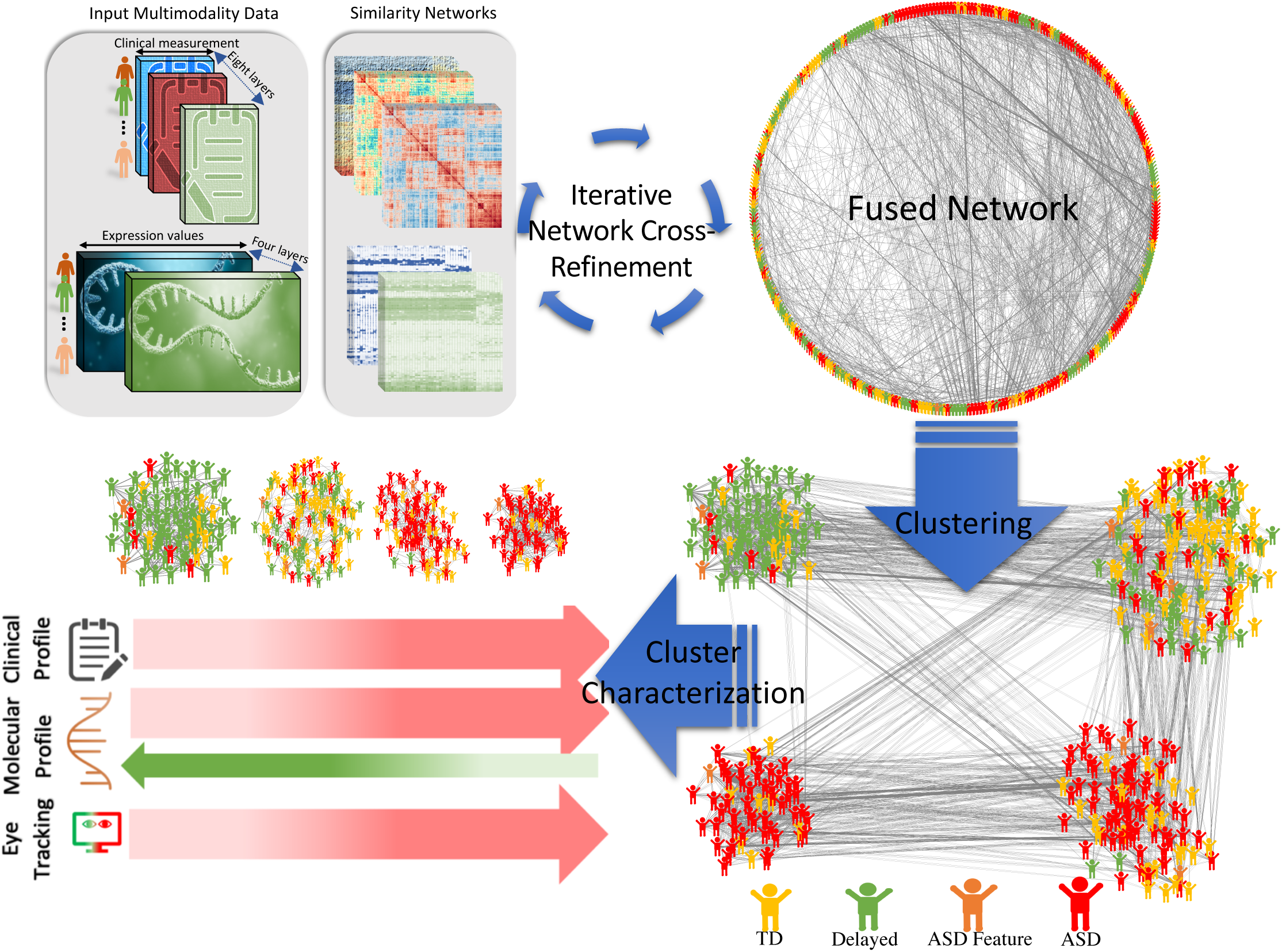
**A.** Schematic view of the workflow used for clustering. First, the 12 patient similarity networks (PSNs) (including eight clinical and four transcriptomics PSNs) were fused using SNF. Then by applying spectral clustering we found the optimal clustering based on the similarity scores in according to 17 different clinical and eye tracking indices that had four different clusters. Among the entire cohort, 42% of toddlers were diagnosed with ASD, 38% with TD, and the remaining with Delay. Cluster 1 primarily consists of toddlers with TD, with a small proportion exhibiting high functioning delayed ASD (14% and 7% respectively). Cluster 2 represents a mixed cluster, with 49% TD, 29% ASD, and 31% Delay diagnoses. Cluster 3 is predominantly comprised of toddlers with ASD, with subtype 1 exhibiting higher ASD symptom severity compared to clusters 1 and 2, but lower severity compared to cluster 4. Cluster 4, identified as ASD subtype 2, comprises 93% ASD diagnoses with no TD toddlers. This cluster exhibits the highest ASD symptom severity and demonstrates a unique eye-tracking pattern.

##### Clinical PSNs

Each PSN was generated by assessing the similarity between subjects based on a single clinical measure. Specifically, we utilized three measures extracted from the Mullen Scales of Early Learning: expressive language (EL), receptive language (RL), and the standardized early learning composite (ELC) score. Additionally, we incorporated five standardized measures from the Vineland Adaptive Behavior Scales, encompassing daily living score, social score, adaptive behavior composite score (ABC), and total motor score. Consequently, we created a total of eight clinical PSNs, each corresponding to one of these clinical measures.

##### Transcriptomic PSNs

The first transcriptomic PSN was constructed based on the similarity of the whole transcriptome. Pearson correlation coefficients (PCC) of the corresponding expression vectors were used to compute the similarity for each pair of subjects. The second transcriptomic PSN was constructed based on the expression of those lncRNAs that were differentially expressed comparing ASD (N= 152) to nonASD (N=211) at p-value<0.05. The third transcriptomic PSN was constructed in a similar way based on sncRNAs. Finally, the last transcriptomic PSN was formed based on the GSVA activity scores associated with the 50 hallmark pathways.

#### 2.6.2 Subtyping

The 12 PSNs were fused using the *similarity network fusion* method implemented in *SNFtool* R package (Wang *et al*., 2014). We applied spectral clustering on the fused network to find the subtypes. For obtaining the optimal clustering we tried different clustering with 3-10 clusters. To score each clustering, first, we took all pairs of candidate clusters and used 16 different clinical indices (including language development, social indices, brain activation, and eye tracking measures) to quantify between-group separation. We used an appropriate statistical test (see the Statistical Analysis and Fig. S1) to assess the difference between each pair of cluster (separation) based on the clinical indices. Then, the proportion of the pairwise cluster differences with a p-value <0.05 was considered as the clustering score. Finally, the clustering with the highest score was selected for defining the subtypes.

### 2.7 Subtyping significance and robustness

Statistical significance of the resulting subtypes was done using ‘sigclust’ R package (Huang *et al*., 2015). To assess the clustering robustness, we used repeated under-sampling on the original data, and the SNF clustering was done on the undersampled dataset using the same parameters. We iteratively and randomly selected subsets of the original data with sizes of 95%, 90%, 80%, 70%, 60%, and 50%. Then using the same procedure, the clustering was done in each iteration. The number of clusters in each clustering was set the same as the optimal number of clusters determined using the original data. The similarity of the obtained clustering in each iteration with the original clustering was assessed using the Jaccard index. The average Jaccard score for all clusters in each iteration was computed as the similarity score between the original clustering and the clustering of the subsampled dataset. This process was iterated 1000 times for each subset size. We used the same data set sizes and did the repeated random partitioning to calculate the background distribution of the Jaccard index in this clustering scenario.

### 2.8 External validation and clinical enrichment of the clusters

We used seven different clinical and eye tracking measures (xSNF) which were not used as input for clustering to validate the obtained clusters. Including three measures showing the ASD symptom severity (see below for more details).

#### Clinical metrics

The total ADOS RRB, and ADOS SA/COSO were used as the two main metrics that measure the ASD symptom severity. These two metrics measures restricted and repetitive behavior, and communication and social of toddlers, respectively. Also, we used the ADOS total which is the sum of the two previous metrics. In addition, we used fine motor (FM) and visual reception (VR) from Mullen Scales of Early Learning.

#### Eye tracking metrics

We used ‘The GeoPref’ eye tracking test which basically assesses the social vs. non-social preference in toddlers (Wen *et al*., 2022). This test consists of two rectangular areas of interest (AOIs) each containing social or non-social (dynamic geometric) video. The total fixation duration within the social AOI was divided by the total fixation duration across the entire video to compute geometric percent fixation (FixationGeo). Also, *((total number of fixations in social AOI) −1) / (total social fixation duration)* was used to calculate saccades/sec in the social AOI. Eye tracking data was collected using the Tobii T120 (Tobii, Stockholm, Sweden; www.tobii.com; 60 Hz sampling rate; 1280 × 1024). We filtered out eye tracking data with poor calibration quality.

### 2.9 Statistical Analysis

Basically, we used ANOVA and ANCOVA for assessing the cluster differences considering each clinical measure (Figure S1). However, according to the different violations from the basic assumptions we used other statistical tests. Shapiro-Wilk and Levene’s test was used for checking the normality and homoscedasticity, respectively. When these tests showed a significant (p<0.05) violation, we used Kruskal-Wallis and Welch tests for assessing the cluster differences, respectively. The R package *rstatix* was used for applying these tests. In cases where both tests showed significant violations, we used quantile-based ANOVA (QANOVA) implemented in the *GFD* package. We also tested the age effect and age-cluster interaction. For those situations with an age effect but not age-cluster interaction we used ANCOVA and age was used as the covariate. Finally, if there were a significant age-cluster interaction (p<0.05) then the final model was selected from the simpler model and the more complex model (including the age-cluster interaction) according to the Bayesian information criteria (BIC). Therefore, if the simpler model had the lower BIC, we didn’t include the interaction term otherwise we included it. In the latter case, we used moderated regression for the cluster comparison in three age bins (mean and mean ± sd). In the former case, we used ANCOVA but we also did moderated regression to check the consistency of the overall interpretation and conclusion. BIC was calculated using the *stats* R package. For the posthoc tests, we used different statistical tests based on the assumption violations. Totally, we used five different posthoc tests for the pairwise cluster comparisons: t-test, t-test with no assumption of equal variance (both implemented in the R package *rstatix*), Wilcoxon-Mann Whitney, robust two-group ANCOVA (implemented in the R package *WRS2*), and estimated marginal means considering age as the covariate (implemented in the R package *emmeans*).

### 2.10 Differentially expressed genes and pathways

For finding the DE genes within different subtypes compared to each other, we applied edgeR on the corresponding normalized and batch effect adjusted read counts. Moreover, to find dysregulated Hallmark pathways, we applied limma to the GSVA activity scores. In both cases, p-values were corrected using the Benjamini-Hochberg method.

### 2.11 Mutations in ASD risk genes

From prior studies, aimed at detecting rare severe mutations in autism and other neurodevelopmental disorder risk genes, we also had single-molecule molecular inversion probes (smMIPs). In those experiments, we did a targeted sequencing on the coding regions for two sets of risk genes.

### 2.12 Brain activation

Functional magnetic resonance imaging (fMRI) data were obtained from 69 subjects. The fMRI task was consistent with protocols used in our prior publications (Redcay and Courchesne, 2008; Eyler, Pierce and Courchesne, 2012; Lombardo *et al*., 2018). Imaging was conducted using a 1.5 Tesla GE MRI scanner (GE High-Definition 1.5 T twin-speed EXCITE scanner) while toddlers were naturally asleep at the University of California, San Diego. Echoplanar imaging was used to acquire functional images (TE = 35ms; TR = 2500ms; flip angle = 90°; matrix size = 64 × 64; resolution = 4 × 4 mm; slice thickness = 4 mm; FOV = 256 mm; 31 slices; 154 volumes).

Preprocessing of functional imaging data, including motion correction, normalization to Talairach space, and smoothing with an 8 mm FWHM Gaussian kernel, was performed using AFNI (Cox, 1996). Brain activation analyses were conducted using the general linear model implemented in SPM8 (http://www.fil.ion.ucl.ac.uk/spm8/).

### 2.13 Outcome patterns and diagnosis stability

Toddlers in this cohort had up to five follow-up visits. In each follow-up visit, all evaluations including ADOS, Mullen Scales of Early Learning, and Vineland Adaptive Behavior Scales were done independently, and finally, a diagnosis was assigned to the subject. We used the sequence of the diagnoses to assess the diagnosis stability within each cluster. In addition, to examine if subjects in different clusters progress differently in terms of symptoms severity, cognitive, language, and social abilities we used a linear mixed model (Bates *et al*., 2015) for each clinical measure of interest. The model has the following form:

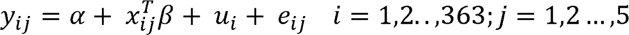

where y_ij_ is a clinical measure of the i^th^ subject (i=1,2,3,…,363) at the j^th^ time point, a is a shared intercept term, x_ij_ is the covariate vector for the fixed effect, β is the coefficient of the fixed effects, u_i_ is the random effect, and *e*_ij_ is an error term (u_i_ ∼ N(0, σ^2^) and *e*_ij_∼ N(0, σ_e_ ^2^)).

## 3. Results

### 3.1 SNF reliably identified 4 clinical-gene expression clusters

We identified 4 clinical-gene expression clusters that vary in clinical scores from high to severely low and gene expression activity from neurotypical to significantly higher than neurotypical, shown schematically in Figure 1. Notice the near complete absence of similarity connectivity between Cluster 4 and Cluster 1 and weak connectivity between Cluster 3 and Cluster 1.

### 3.2 Clustering validation

For iSNF clinical variables and the xSNF variables (external validators) including ADOS, visual reception and eye tracking measures, statistical tests (ANOVA, ANCOVA, and QANOVA) showed highly significant differences between clusters (Figure S2 and S3). Also, figure 2A shows the distribution of all iSNF and xSNF clinical measures. Additionally, we assessed clustering robustness by repeatedly under-sampling the original data and applying SNF clustering with the same parameters (Figure 2B). Subsets of 95%, 90%, 80%, 70%, 60%, and 50% of the original data were randomly selected (see the Methods for more details). This shows highly significant robustness of the difference between the Jaccard index values in random clustering and optimal clustering (p<0.001). Sigclust also showed that the clustering is highly significant (p-value < 0.001).

**Figure 2:**
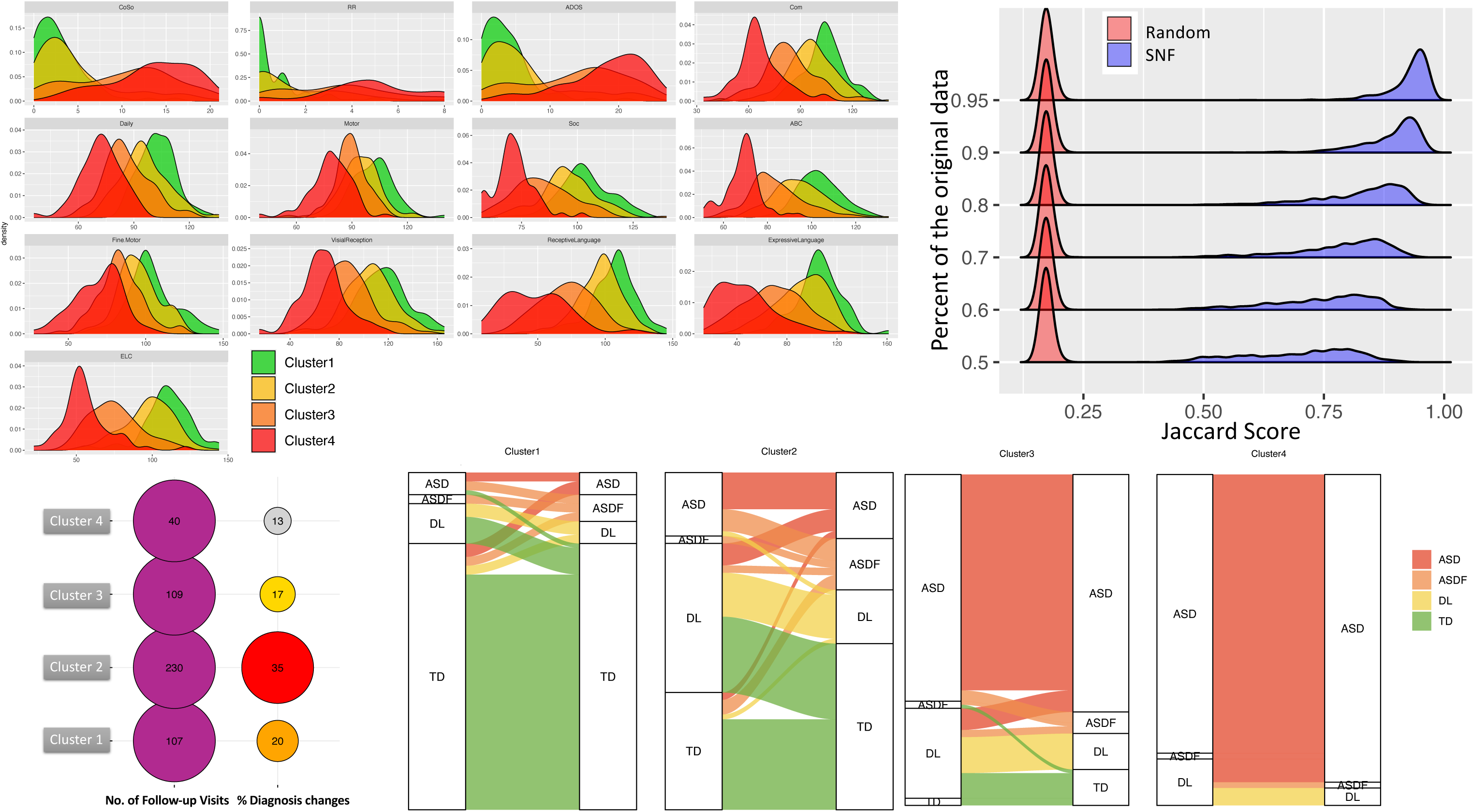
A. The distribution of 13 clinical measures, comprising 8 iSNF measures used to construct the clinical patient similarity networks (PSNs), which were used as SNF input, and 5 xSNF measures, which were not used for clustering but for validating the obtained clusters. The iSNF clinical measures include expressive language (EL), receptive language (RL), early learning composite (ELC), daily living, social score, adaptive behavior composite (ABC), total motor score, and 5 xSNF measures: ADOS RRB, ADOS SA/COSO, ADOS total (ASD symptom severity), fine motor (FM), and visual reception (VR). All clinical measures, both internal (iSNF) and external (xSNF), exhibit distinct distribution patterns across clusters (see also Figure S2 and S3). **B.** Clustering robustness. We assessed clustering robustness by repeatedly undersampling the original data and applying SNF clustering with the same parameters. Subsets of 95%, 90%, 80%, 70%, 60%, and 50% of the original data were randomly selected. Clustering was performed with the optimal number of clusters determined from the original data, and the Jaccard index measured the similarity. This process was repeated 1000 times for each subset size, and the resulting distribution of Jaccard indices was plotted. The y-axis represents different percentages of the original data randomly selected, while the x-axis shows the averaged Jaccard index distribution. **C.** Diagnosis stability in each cluster. The left column displays the total number of follow-up visits within each cluster. The right column illustrates the percentage of diagnosis changes observed between two consecutive visits. For instance, within cluster 2, there were a total of 230 follow-up visits, with diagnosis changes occurring in 35% (81 out of 230) of these visits. **D:** Diagnosis changes categorized by diagnosis group per cluster. The first column shows the diagnosis at the first visit and the second column shows the diagnosis at the last visit. This panel shows the diagnosis change trends within each cluster. For example, in cluster4: all ASD subjects stayed in the same diagnosis group, also four delayed and one ASD feature toddlers changed to ASD, and finally the diagnosis for one toddler changed from delayed to ASD features. On the other hand, in cluster1, 88% of the TDs stayed in the same diagnosis group while 7 toddlers transitioned from other diagnosis to TD (6 from LD to TD and 1 from ASD to TD).

### 3.3 Four main clinical-gene expression clusters

Using SNF to integrate the clinical and molecular data from n = 363 ASD, TD and DL toddlers, four clusters were found (Figure 1): 93% of Cluster 4 were ASD (n=53) and 79% of Cluster 1 were TD. Also, Cluster 3 was 72% moderately affected ASD and Cluster 2 was a “mixed” Cluster of better able ASD (n=28), lower ability TD (n=67) and Delayed (n=43) toddlers (Table 2).

**Table 2:**
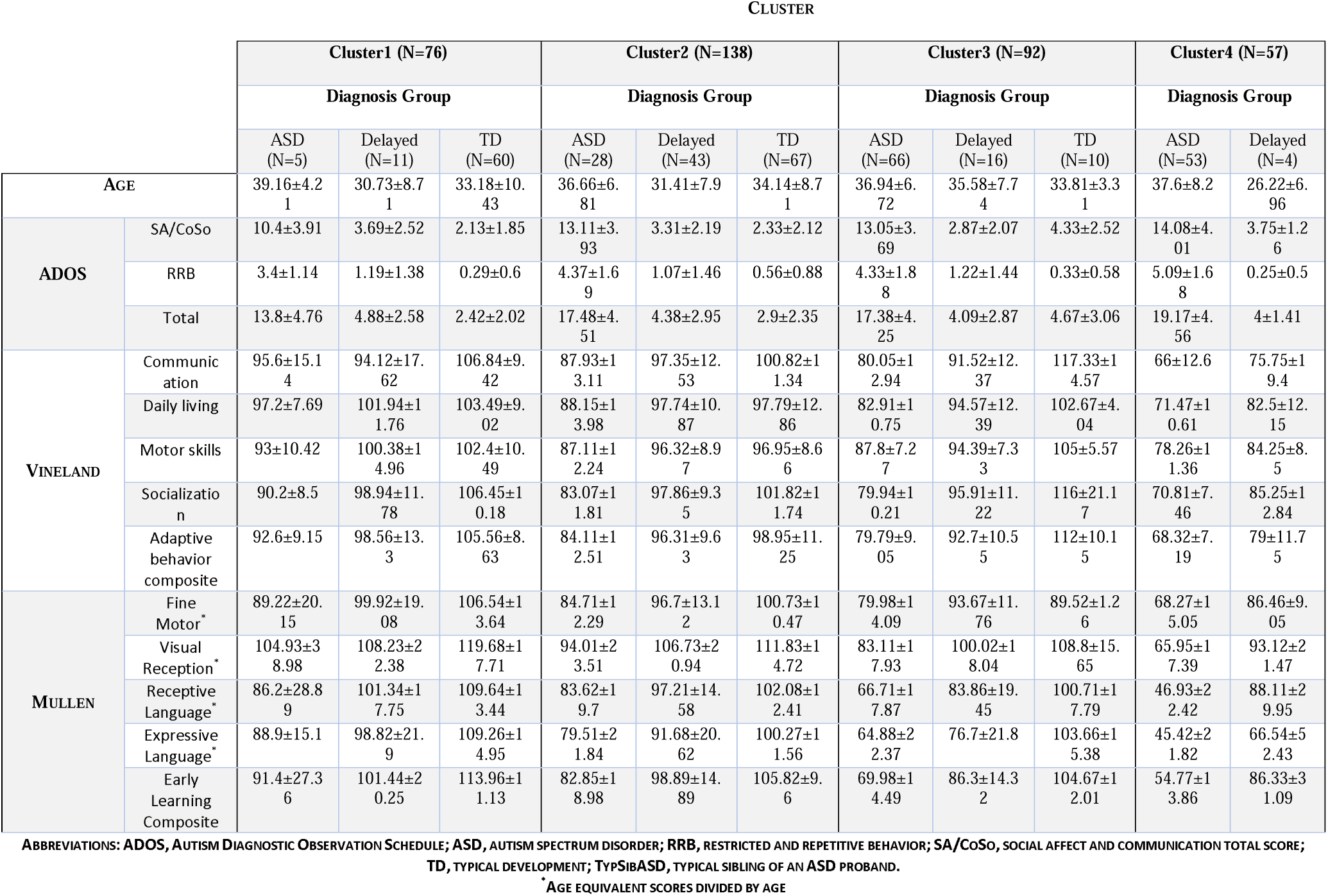
Clinical Characteristics of Toddlers Used in This Study Stratified by Cluster and Diagnosis Group at the follow-up visit.

#### High diagnostic stability of profound autism

There were 486 diagnostic and psychometric longitudinal follow-up visits for subjects in the four clusters (Figure 2C), with 125 resulting in a change in diagnostic classification at the most recent diagnostic testing compared to the first testing. This relatively small change is consistent with the recent Pierce et al report on diagnostic stability in N=1,269 ASD, typical and non-ASD delayed toddlers (Pierce *et al*., 2019). Figure 2D is a Sankey diagram of these longitudinal changes for ASD, TD, delayed and ASD features separately from delayed and ASD.

Toddlers in the profound autism Cluster 4 had 100% diagnostic stability: that is, no profound autism Cluster 4 toddler changed to a different diagnosis at the most recent diagnostic testing (later-age) compared to the first visit. Cluster 3 was also stable (81%) with only three ASD toddlers re-diagnosed as ASD features (Figure 2C and 2D) and one ASD toddler re-diagnosed as TD. TD Cluster 1 was also relatively stable, but as previously reported by Pierce *et al*. (Pierce *et al*., 2019), a small percentage of TD change to a high or mild ability ASD diagnosis at follow-up.

### 3.4 Diagnostic and subtype differences in clinical scores

Table 2 shows clinical scores for each of the four clusters at their follow-up visit as well as for the ASD, TD and Delayed subjects separately within each cluster. ASD toddlers in Cluster 4 (93% ASD) have clinical scores consistent with profound autism (Table 2, Figure 2A and Figure S2). They have high social symptom severity and low receptive and expressive language and low overall cognitive IQ, with mean scores below −3 standard deviations (SDs) (Table 2; Figure 2A and Figure S2). In sharp contrast are ASD toddlers with mild clinical features who are in TD Cluster 1 (79% TD) and Cluster 2, which is a mix of mild ASD, low TD and delayed toddlers. ASD patients in Cluster 2 have psychometric scores generally within the neurotypical range, and the very small subset in Cluster 1 are the highest ability ASD toddlers among the mild autism range; potentially among these few are individuals who may eventually have an “optimal” outcome. Nearly exactly in between are ASD toddlers with moderate social, language and cognitive difficulties, not nearly as severe as the profound or as neurotypical as the mild toddlers.

### 3.5 ASD subtype separation validation and comparisons with TD and Delayed

Statistically significant differences were found between ASD patients in Clusters 4 vs 3 vs 2+1. Comparisons between ASD patients in Cluster 4 and Cluster 3 vs all TD subjects were also highly significant. Comparisons between ASD patients in Clusters 1+2 vs all TD subjects showed significant differences in all clinical measurements.

Interestingly, despite clustering together, ASD and Delayed toddlers within Clusters 4 and 3 were also quantitatively and statistically distinguishable. It is important however to emphasize the Delayed toddlers are a mix of different types of delay such as late talkers who improved or declined in expressive language with age, motor delay, some features of ASD and other developmental differences (eg, ADHD).

In sum, SNF differentially identified a highly stable and distinct profound autism subtype that is about 30% of the ASD sample as well as better ability subtypes of ASD based on clinical and in vivo gene expression data.

### 3.6. Profound autism subtype-specific and ASD group-nonspecific pathway dysregulations

Comparison of Hallmark pathway activity scores of all ASD toddlers versus all TD toddlers found 17 significantly upregulated Hallmark pathways (Figure 3). Four of these 17 were significantly dysregulated in the profound autism subtype: estrogen response early and late, bile acid metabolism and heme metabolism.

**Figure 3:**
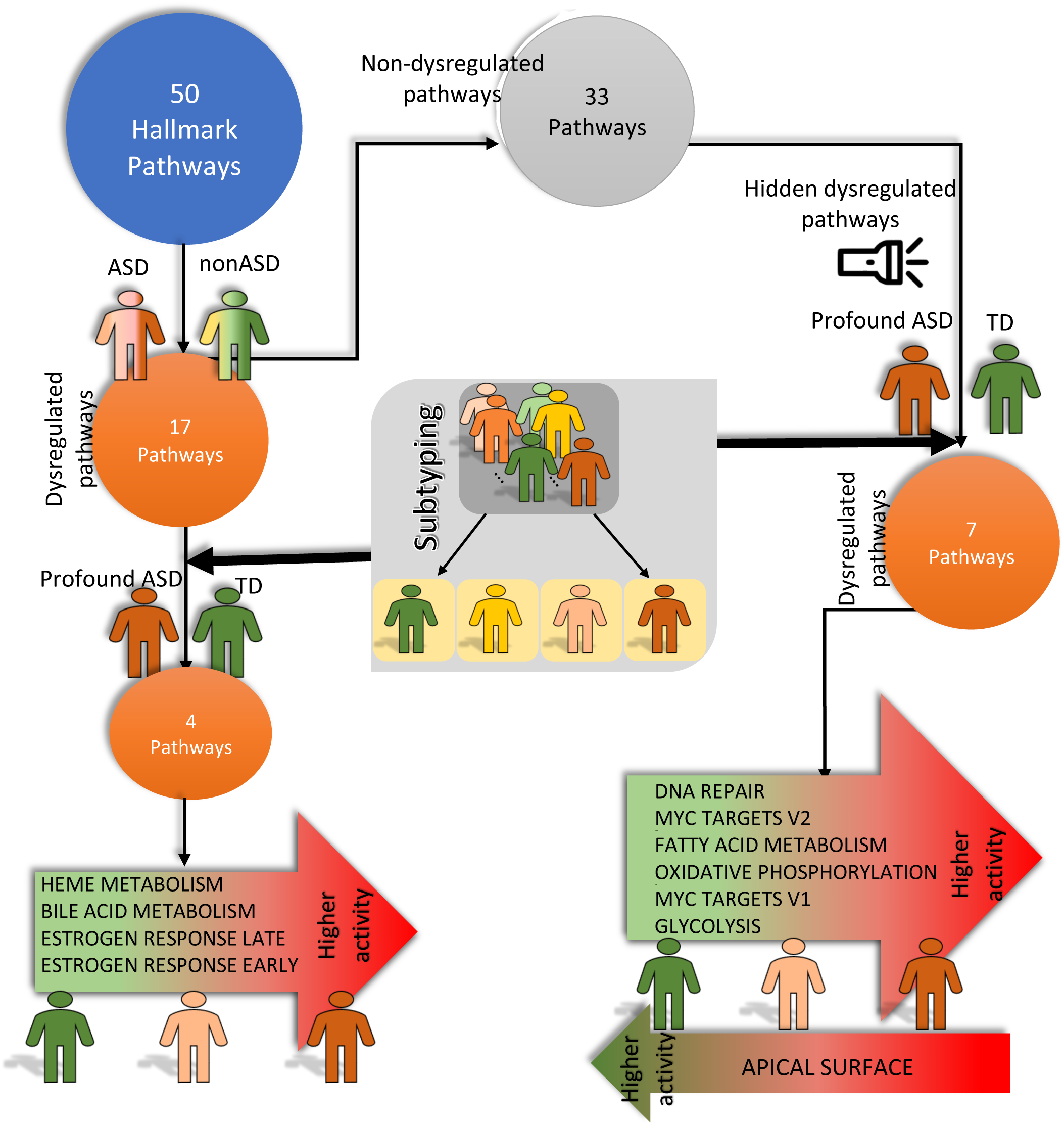
Pathway dysregulation in ASD and ASD subtypes. We used gene set variation analysis (GSVA) to assess the activity of 50 hallmark pathways from the Molecular Signatures Database (MSigDB). Results revealed increased activity of most hallmark pathways in ASD compared to TD (45/50 pathways). Notably (left path), 17 pathways were significantly upregulated in ASD (FDR q-value <= 0.1). To explore cluster-specific differences, we focused on these 17 pathways. Excluding cluster2 with mixed diagnoses, we found four pathways (HEME METABOLISM, BILE ACID METABOLISM, ESTROGEN RESPONSE LATE, ESTROGEN RESPONSE EARLY) dysregulated between TD and the profound ASD subtype (Clusters1 vs Clusters1). Heme metabolism was the only significantly dysregulated pathway between two ASD subtypes (Clusters 3 & 4) and TD. Considering the heterogeneity in ASD, it is likely to miss some transcriptomics dysregulation when ASD is compared to TD as a whole group. To find potential hidden pathway dysregulations that are hidden in diagnosis-level comparisons, we further analyzed the 33 non-dysregulated pathways and identified seven pathways with differential activity between TD and profound ASD subtype (right path). Six pathways showed higher activity in the profound ASD subtype (Cluster 4), while one (APICAL SURFACE) displayed the opposite pattern. Overall, 11 hallmark pathways exhibited a continuum of dysregulation, aligning with the phenotypic severity of ASD subtypes.

Next, we tested for subtype-specific dysregulations and found 6 pathways differentially overactive and 1 underactive in profound autism as compared to other ASD subtypes. Overactive pathways were Myc targets (v1 and v2), oxidative phosphorylation, glycolysis, DNA damage repair, and fatty acid metabolism, and underactive was apical surface (Figure 3).

As shown in Figure 4A, these eleven pathways play key roles during embryogenesis including cell cycle progression, proliferation, asymmetrical division, apical polarization, differentiation, neurogenesis, hedgehog signaling, and Wnt signaling and neural growth during embryogenesis. Thus, toddlers with profound autism have dysregulations in multiple embryonic gene pathways, including processes involved in brain cortical organoid (BCO) enlargement and accelerated growth and neurogenesis in profound autism toddlers (Courchesne et al., 2024).

**Figure 4:**
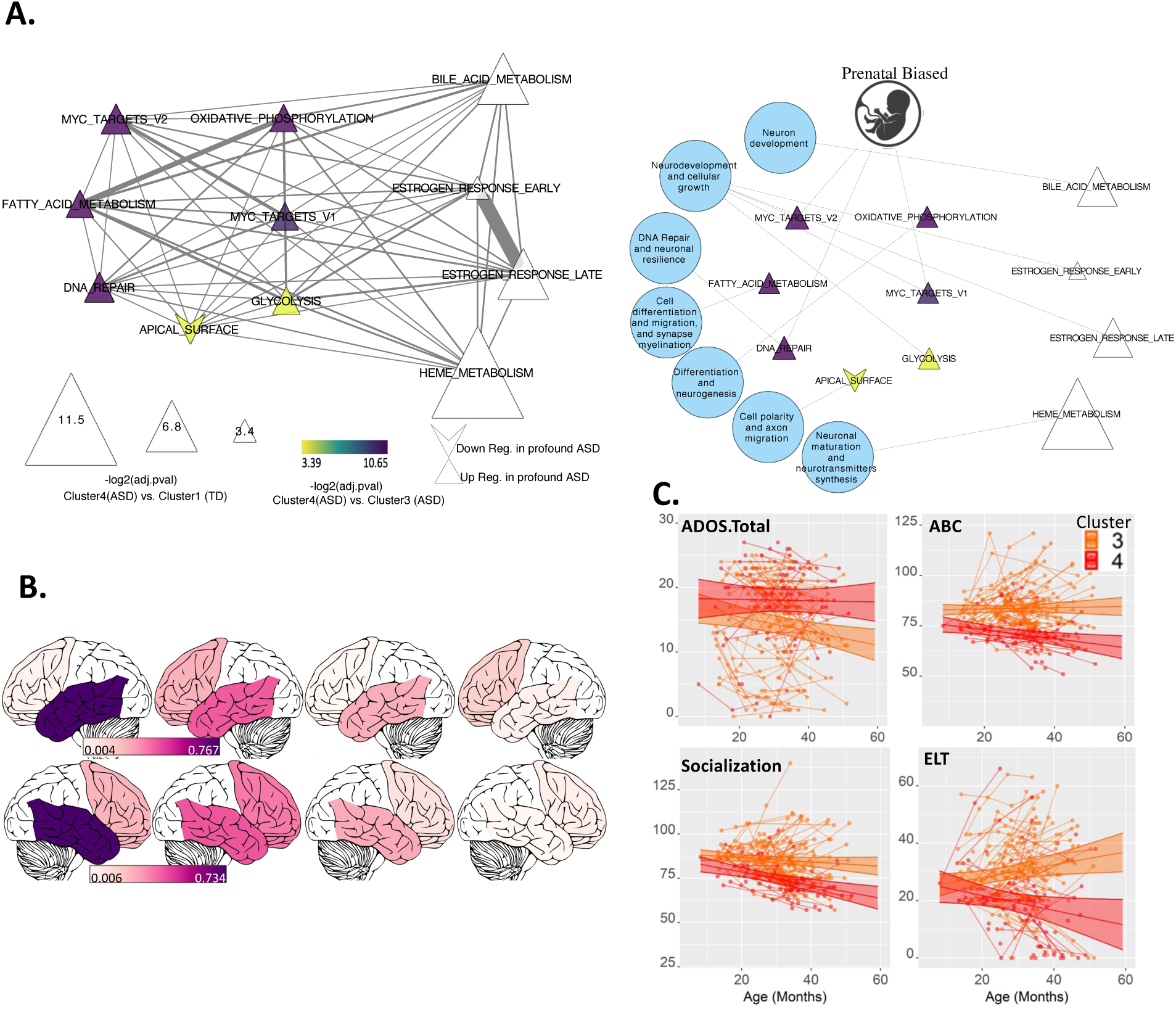
A. Network visualization depicting the interplay among the 11 dysregulated pathways. Left Network: edge thickness represents the number of shared genes between pathway pairs, while node size corresponds to the −log(adjusted p-value) when comparing the ASD profound subtype to the TD group within cluster1. Colored nodes highlight significant dysregulation (adjusted p-value <= 0.1) between ASD subtypes (cluster 3 and cluster 4), with color intensity reflecting the −log(adjusted p-value) when comparing the ASD profound subtype to the mild ASD subtype in cluster3. Additionally, node shape indicates the direction of dysregulation, with upright triangles representing upregulation in the ASD compared to TD and upregulation in ASD profound subtype to the mild ASD subtype and inverted triangles representing downregulation. B. Brain activation based on fMRI data from 69 toddlers during natural sleep. Clusters 3 and 4 exhibited reduced temporal lobe activation compared to cluster 1. No significant differences were found between profound and mild ASD subtypes, likely due to the small sample size. Note: Regions are visualized for simplicity and are not exactly the same as those used as ROIs. C. Longitudinal assessment of outcome disparities between two ASD clusters, conducted through a mixed effect model analysis across four primary clinical measures. Notably, three clinical metrics exhibited significant differences between the two clusters (p-value<5E-2). Of particular interest is the trajectory observed in the ELC measure: individuals in cluster 3 exhibit improvement with age, whereas those in cluster 4 display a contrasting trend (p-value = 2E-03). A similar pattern is evident in the ABC metric (p-value = 1.6E-02). In terms of socialization, both clusters show a decline, yet cluster 3 exhibits a shallower slope (p-value = 3.7E-02). Furthermore, distinct changes in symptom severity are discernible between the clusters: while cluster 4 demonstrates relative stability, cluster 3 experiences a worsening trend (although the difference is less pronounced; p-value = 1.1E-01). Right network: the same dysregulated pathways which are linked to the ASD-related biological processes. Also, three pathways (MYC TARGETS V1 and V2, and OXIDATIVE PHOSPHORYLATION) are significantly over active prenatally according to gene expression data from developing brain (Brainpan database).

MIR3648-1, a human-specific miRNA, was the only dysregulated sncRNA. It was upregulated in the profound autism subtype vs moderate ASD Cluster 3 (log fold change: 2.3; FDR q-value 0.002). It is broadly expressed in cortex, cerebellum, and blood (Supplementary Figure S4 A); promotes cell proliferation (Rashid et al., 2017; Xing, 2019), and plays a role in the differentiation of mesenchymal stem cells (Min et al., 2019). Three lncRNAs were upregulated in profound autism vs moderate ASD Cluster 3: FP236383.4, FP236383.5, and FP671120.7, while only one was significantly downregulated, AC130304.1. All of these three LNCs are broadly expressed (Supplementary Figure S4 B-C)

### 3.7 Rates of SFARI Level 1 and 2 gene mutations are similar in ASD and TD

The only biological metric we did not find related to ASD social clinical-gene expression subtypes was DNA risk gene mutations. The ASD group was not enriched in SFARI Level 1 and 2 risk gene mutations compared to nonASD toddlers. Interestingly, the ASD group was slightly under-enriched (hypergeometric P-value>0.25, under-enriched 1.26 fold) and nonASD group was slightly over-enriched (hypergeometric P-value>0.25, over-enriched 1.12 fold). Also, none of the ASD subtypes nor clusters were significantly under- or over-enriched in SFARI Level 1 and 2 risk gene mutations. The most frequent mutation was missense mutation in nonASD subjects in cluster2.

### 3.8 Reduced Temporal Lobe Activation in Clusters 3 and 4 Compared

fMRI data from 69 toddlers were analyzed to examine brain activation during natural sleep (Figure 4.B). Significant differences in brain activation were observed between clusters in both the left and right temporal lobes. Pairwise comparisons revealed a significant reduction in temporal lobe activation in clusters 3 and 4 compared to cluster 1 (all p-values < 0.05). This suggests that brain activation patterns in the temporal regions vary significantly across these clusters, with clusters 3 and 4 showing notably less activation than cluster 1. No significant differences were observed between the profound and mild ASD subtypes, likely due to the small sample size.

### 3.9 ASD Subtype Clinical Outcome Trajectories

Taking advantage of the clinical longitudinal subject data (Figure 2C), we assessed outcome differences between profound and moderate ASD subtypes using a mixed effect model in four main clinical measures (see Figure 4C). Three clinical measures showed differences between the two ASD subtypes (at p-value<0.05). Figure 4C shows the trajectory for all four main clinical metrics for the profound and moderate autism subtypes. Toddlers in the profound autism subtype (in red) remained high in social symptom severity across age, but toddlers in the moderate ASD subtype (in orange) showed improvement in ADOS social symptoms (p-value = 0.1). On the Vineland Socialization and overall Adaptive Behavior Composite, profound autism declined significantly, but the moderate ASD did not decline (p-value< 0.04). Toddlers with moderate ASD displayed substantial improvement in expressive language with age, toddlers with profound autism declined substantially

## 4. Discussion

To objectively identify toddlers with profound autism and those with milder ASD phenotypes, we deployed SNF subtyping, but excluded ADOS scores so that this diagnostic tool cannot drive subtyping (Figure 1). With SNF, we integrated molecular and clinical (Mullen, Vineland) data in a large ASD sample representative of the general population in San Diego County and identified three main ASD molecular-clinical subtypes, as well as a tiny subset among the mild ASD subtype who may be provisionally described as “optimal” autism. One subtype is profound autism that was categorically different from moderate and mild ASD and control subtypes. This profound autism subtype is in a cluster composed of 93% ASD toddlers with low social, language, cognitive scores and severe social symptoms; this clinical profile is consistent with profound ASD (Lord *et al*., 2022).

Using analyses of MSigDB Hallmark pathways, we discovered molecular differences specific to profound autism. These differences occurred in 7 Hallmark gene pathways known to govern progression of multiple stages in embryonic brain development from cell proliferation to differentiation of cortical cells to neurons (Figure 4A). This result is consistent with our theory of ASD (Courchesne *et al*., 2019; Courchesne, Gazestani and Lewis, 2020) and, importantly, our new evidence from brain cortical organoids (BCO) that profound autism begins embryonically with extreme embryonic BCO enlargement, growth rate and accelerated neurogenesis. In that study as in the present one, those profound autism toddlers had severe social symptoms, reduced social attention, severely reduced language abilities, and atypical growth of social, language and sensory cortices (Courchesne *et al*., 2024).

Five of these 7 Hallmark pathways are directly or interactively involved in cell cycle, proliferation, differentiation, neurogenesis, DNA replication during cell division, and growth and energy metabolic processes and the progression of one stage to the next (Figure 4A). For example, Myc pathways, glycolysis, and oxidative phosphorylation are “master regulators” of cell cycle, proliferation metabolism and differentiation as well as DNA replication and damage repair (Kuwahara *et al*., 2010a; Mainwaring, Bhatia and Kenney, 2010; Zheng *et al*., 2016a). In ASD, overly rapid and disorganized cell cycle, excess proliferation, overabundance of cortical neurons and early brain overgrowth have been reported, notably in ASD toddlers and cell models derived from them (Courchesne *et al*., 2011; Pramparo, Lombardo, *et al*., 2015; Pramparo, Pierce, *et al*., 2015; Marchetto *et al*., 2017; Lombardo *et al*., 2021; Bao *et al*., 2023). We hypothesize excess Myc upregulation in ASD toddlers may be related to embryonic BCO accelerated growth, accelerated cell cycle progression, excess proliferation, accelerated neurogenesis, and prenatal BCO overgrowth that is correlated with social symptom severity (Courchesne *et al*., 2024), and with early postnatal brain overgrowth, atypical growth of social and language cortex and, ultimately, reduced social neural activity in temporal cortex (Figure 4B).

These subtype-specific overactive Hallmark gene pathways in profound autism are regulated by upstream signaling of RAS-ERK, PI3K/AKT/GSK-3 and Wnt/β-catenin signaling which are also overactive in ASD toddlers (Gazestani *et al*., 2019); overactive in ASD embryonic neural progenitor cells and neurons derived from ASD toddler; and overactive in ASD toddlers with the most severe ASD social symptoms (Gazestani *et al*., 2019). Neuronal development is highly energy-demanding, and the transition from aerobic glycolysis in proliferating cells to oxidative phosphorylation in differentiated neurons, is another example of the interplay among these Hallmark pathways during which Myc normally downregulates (Zheng *et al*., 2016b; Rumpf, Sanal and Marzano, 2023). Thus, this set of dysregulated Hallmark pathways are tightly interrelated in crucial neurobiological programs in embryogenesis.

Myc is directly regulated by the autism-relevant β-catenin/Tcf transcription complex and mediates functions of the Wnt signaling pathway, stimulating neocortical neural progenitor cell (NPC) proliferation and differentiation (Kuwahara *et al*., 2010b). Estrogen response pathways also enhance the expression of Wnt3 the Wnt/β-catenin signaling pathway, leading to its activation (Zhang *et al*., 2008). Consistent with this, we found the upregulation of Wnt/β-catenin using microarray data in another ASD toddler cohort (Gazestani *et al*., 2019). The degree of dysregulation in these pathways in that study were linked to ASD social symptoms severity, with more pronounced dysregulation corresponding to more severe manifestations of ASD. More broadly, estrogen response pathways play a significant role in the development and function of the nervous system, particularly in shaping the structure and plasticity of the brain (McCarthy, 2008).

Another profound autism subtype-specific Hallmark pathway is the DNA repair pathway. This pathway is critical for ensuring genomic integrity during embryonic development, as it is believed that major DNA repair pathways are functional in embryos (Khokhlova *et al*., 2020). Previously, dysregulation of DNA damage repair functions has been reported postmortem in ASD at older ages (Chow *et al*., 2012) and suggested to underlie somatic mosaicism and focal cortical dysplasia (Stoner *et al*., 2014). Moreover, the DNA repair process is intimately connected with cellular metabolism. Metabolic processes affect DNA repair, and conversely, DNA damage can induce metabolic functional activity (Turgeon, Perry and Poulogiannis, 2018). DNA damage response activation can prompt an upsurge in nucleotide synthesis and the metabolic processing of glucose. Its varied activity across the ASD subtypes suggests a role in responding to developmental stress and neuronal damage. The degree of dysregulation in this pathway could reflect the resilience of neuronal populations in different ASD subtypes. The above-mentioned pathways could be linked to on-average larger brain size in severe ASD subjects and to lower brain activity and function.

Lastly, this tight constellation of dysregulated embryogenesis-important pathways in profound ASD may also explain why they were the only ASD patient subtype to not have age-related clinical improvement (Figure 4C).

In addition to these embryonic-important pathways in the profound ASD subtype, 17 other Hallmark pathways were significantly upregulated across ASD subtypes as compared with nonASD toddlers; no pathway was significantly downregulated in ASD as compared to TD toddlers (Figure 3). Among these were metabolic pathways crucial for brain function that show varying degrees of activity across the ASD subtypes. These include heme metabolism and bile acid metabolism pathways. Fatty acid metabolism is crucial for the biosynthesis of cell constituents, such as lipids, that are fundamental for developmental processes like synapse formation and elimination, as well as myelination (Nutr and Steiner, 2019). Additionally, the role of docosahexaenoic acid (DHA), a major product of fatty acid metabolism, is significant in neurogenesis. The expression of fatty acid-binding proteins (B-FABP) during development parallels early neuronal differentiation and is thought to play a crucial role in early neurogenesis or neuronal migration (Innis, 2007). This pathway’s influence on synapse formation, neurogenesis, and cognitive functions suggests that its altered activity in different ASD subtypes might contribute to the observed variability in symptom severity, cognitive abilities, and sensory processing. Understanding the modulation of this pathway in ASD could provide critical insights for developing targeted interventions aimed at improving neurodevelopmental outcomes in individuals with ASD. The heme metabolism pathway is critical for various aspects of brain development and function. During early postnatal development, iron, a crucial component of the heme group, is employed extensively for mitochondriogenesis, neuronal maturation, synthesis of neurotransmitters, and myelination (Chang *et al*., 2005; Ozsoy, 2020). The bile acid metabolism pathway holds a significant position in neural development due to its role in the management of cholesterol, an essential component for the development of neurons and neuroglia. Notably, a substantial portion of the body’s cholesterol, nearly 25%, is located within the brain, underscoring its critical function in neural structure and function (Grant and Demorrow, 2020). This pathway’s role extends to the synthesis of bile acids, which differ in Alzheimer’s Disease (AD) compared to cognitively normal individuals, as revealed by metabolic network analyses of post-mortem brain samples (Baloni *et al*., 2020). Furthermore, the metabolism of bile acids is intricately linked with cognitive functions. Elevated levels of primary and secondary bile acids have been observed in AD samples relative to controls, along with higher serum levels of taurine in AD patients (MahmoudianDehkordi *et al*., 2019). Some recent studies have postulated that alterations in the gut microbiota, with consequent changes in serum and brain bile acid profiles, may be mechanistically involved in the development of diseases related to cognitive dysfunction, such as mild cognitive impairment (MCI), AD, developmental coordination disorder (DCD), and vascular dementia (VD)(Weng *et al*., 2022). The transformation of bile acids by the gut microbiota is also implicated as a contributing factor in the development and progression of AD and Huntington’s disease (HD)(Jia *et al*., 2020). Collectively, these pathways are essential for synaptic development, neurogenesis, energy production, and neurotransmitter synthesis. Their dysregulation, particularly the higher activity in both ASD subtypes compared to TDs and lower activity in less severe ASD subtype (cluster 3) than severe ASD subtype (cluster 4), may correlate with specific ASD phenotypes, such as cognitive and developmental delays.

Unexpected was that SFARI level 1 and 2 ASD gene mutations were similarly common in typical, delayed and ASD toddlers (Supplementary Figure S6). Moreover, the profound ASD subtype did not have the highest rate of SFARI level 1 and 2 ASD mutations among patients (Supplementary Figure S7). Further, ASD gene mutations were not more common in subtypes with the most Hallmark pathway dysregulations. Instead, they were as common in subtypes with more as well as subtypes with few dysregulated Hallmark pathways. These results are generally consistent with our recent work showing SFARI Level 1 and 2 ASD gene mutations are equally common in typical and ASD toddlers (Bao *et al*., 2023). In that study, SFARI Level 1 and 2 ASD risk genes were not related to diagnostic or psychometric symptom severity in either ASD or typical toddlers and had no detectable impact on expression in genes that are accurate ASD diagnostic markers. In these studies, then, there is no evidence that the SFARI Level 1 and 2 ASD gene mutations detected in our ASD, TD and delayed toddlers have clinical, diagnostic, or functional genomic significance specific to ASD. This raises the question of which SFARI Level 1 and 2 genes do and which not have functional significance for the development of ASD.

This work makes a major advance in fundamental knowledge of ASD molecular subtypes from toddler blood and their close relationship to ASD variation in social symptoms, developmental trajectories, social attention, and social neural function, and is built on a foundation of previous work showing dysregulated developmental gene pathways and modules identified in ASD blood gene expression are correlated with social symptoms, language ability, social neural functioning, and cortical patterning and provide accurate early-age diagnostic signatures. While the present work is remarkable in advancing fundamental knowledge about subtypes of the molecular beginnings of ASD, perhaps even more remarkable is that the field has largely ignored transcriptomics-based approach in favor of studies of risk gene mutations that, in this and other studies, provide no useful information on subtype-specificity or on symptom, brain and behavior heterogeneity in idiopathic ASD at early ages. To date, risk gene mutations provide little insight into the profound autism subtype. This work, therefore, highlights that to gain much deeper foundational knowledge of profound, moderate and mild ASD at the beginning, a wide array of social-relevant modalities must be collected and integrated within-toddler including embryonic model, in vivo gene expression, social fMRI, social neuroanatomy, social attention, social and language ability, and social symptoms. Lastly, so long as ASD treatment studies fail to be aware of multi-modality social subtypes at baseline, designs at the overall group level with no knowledge of the admixtures of subtypes being treated, will not find reproducible treatment effects, nor maximize treatment-specific needs of toddlers in each subtype.

## Data Availability

All data produced in the present study are available upon reasonable request to the authors

## Acknowledgements

The authors sincerely thank the parents and children in San Diego who participated in our research, without whom this would not be possible. We are also fortunate to work with wonderful pediatricians and family practice physicians spanning a range of medical groups including UCSD, Sharp Rees-Stealy, Scripps, Rady-Children’s Primary Care Medical Group, Chula Vista Pediatrics, Graybill Medical Group, Grossmont Paediatrics, Linda Vista Health Care Center, Mills Pediatrics, North County Health Services, San Diego Family Care, and Sea Breeze Pediatrics. We are grateful for their support. This work was supported by NIDCD grant 1R01DC016385 awarded to Eric Courchesne and Karen Pierce; and NIMH grants R01MH118879 and R01MH104446 awarded to Karen Pierce.

## Author contributions

J.Z. and E.C. conceived the idea and designed the study. K.P., L.L. and E.C. collected the data. J.Z. performed all analyses. E.C., N.E.L. and K.P. obtained grant funding. J.Z. and E.C. wrote the manuscript. All authors contributed to editing the manuscript.

## Competing Interests

The authors declare no competing interests.

## Data Code availability

The code for all the analysis can be found at: https://github.com/ACE-UCSD/ASD_Subtyping_TransciptomicsClinical. The datasets generated during and/or analyzed during the current study are available from the corresponding authors on reasonable request.

## Notes

### Competing Interest Statement

The authors have declared no competing interest.

### Author Declarations

The Institutional Review Board approved this study at the University of California, San Diego.

### Summary of Updates

New Authors Have been added

## Reference

1. Ansel, A., et al. (2017) ‘Variation in Gene Expression in Autism Spectrum Disorders: An Extensive Review of Transcriptomic Studies’, Frontiers in neuroscience, 10(JAN). Available at: 10.3389/FNINS.2016.00601.

2. Bai, D., et al. (2019) ‘Association of Genetic and Environmental Factors With Autism in a 5-Country Cohort’, JAMA psychiatry, 76(10), pp. 1035–1043. Available at: 10.1001/JAMAPSYCHIATRY.2019.1411.

3. Baloni, P., et al. (2020) ‘Metabolic Network Analysis Reveals Altered Bile Acid Synthesis and Metabolism in Alzheimer’s Disease’, Cell Reports Medicine, 1(8), p. 100138. Available at: 10.1016/J.XCRM.2020.100138/ATTACHMENT/112351A6-707E-4C55-80E6-264D6A31B9FC/MMC14.XLSX.

4. Bao, B., et al. (2023) ‘A predictive ensemble classifier for the gene expression diagnosis of ASD at ages 1 to 4 years’, Molecular psychiatry, 28(2), pp. 822–833. Available at: 10.1038/S41380-022-01826-X.

5. Bates, D., et al. (2015) ‘Fitting Linear Mixed-Effects Models Using lme4’, Journal of Statistical Software, 67(1), pp. 1–48. Available at: 10.18637/JSS.V067.I01.

6. Chang, E.F., et al. (2005) ‘Heme regulation in traumatic brain injury: Relevance to the adult and developing brain’, Journal of Cerebral Blood Flow and Metabolism, 25(11), pp. 1401–1417. Available at: 10.1038/SJ.JCBFM.9600147/ASSET/IMAGES/LARGE/10.1038_SJ.JCBFM.9600147-FIG8.JPEG.

7. Chiang, A.W.T., et al. (2021) ‘Optimal balancing of clinical factors in large scale clinical RNA-Seq studies’, bioRxiv, p. 2021.06.30.450639. Available at: 10.1101/2021.06.30.450639.

8. Ch’ng, C., et al. (2015) ‘Meta-Analysis of Gene Expression in Autism Spectrum Disorder’, Autism research : official journal of the International Society for Autism Research, 8(5), pp. 593–608. Available at: 10.1002/AUR.1475.

9. Chow, M.L., et al. (2012) ‘Age-dependent brain gene expression and copy number anomalies in autism suggest distinct pathological processes at young versus mature ages’, PLoS genetics, 8(3). Available at: 10.1371/JOURNAL.PGEN.1002592.

10. Courchesne, E., et al. (2011) ‘Neuron number and size in prefrontal cortex of children with autism’, JAMA, 306(18), pp. 2001–2010. Available at: 10.1001/JAMA.2011.1638.

11. Courchesne, E. et al. (2019) ‘The ASD Living Biology: from cell proliferation to clinical phenotype’, Molecular psychiatry, 24(1), pp. 88–107. Available at: 10.1038/S41380-018-0056-Y.

12. Courchesne, E., et al. (2024) ‘Embryonic origin of two ASD subtypes of social symptom severity: the larger the brain cortical organoid size, the more severe the social symptoms’, Molecular Autism, 15(1), pp. 1–16. Available at: 10.1186/S13229-024-00602-8/FIGURES/5.

13. Courchesne, E., Gazestani, V.H. and Lewis, N.E. (2020) ‘Prenatal Origins of ASD: The When, What, and How of ASD Development’, Trends in neurosciences, 43(5), pp. 326–342. Available at: 10.1016/J.TINS.2020.03.005.

14. Cox, R.W. (1996) ‘AFNI: Software for analysis and visualization of functional magnetic resonance neuroimages’, Computers and Biomedical Research, 29(3), pp. 162–173. Available at: 10.1006/cbmr.1996.0014.

15. Diaz-Beltran, L., Esteban, F.J. and Wall, D.P. (2016) ‘A common molecular signature in ASD gene expression: following Root 66 to autism’, Translational psychiatry, 6(1). Available at: 10.1038/TP.2015.112.

16. Enstrom, A.M., et al. (2009) ‘Altered gene expression and function of peripheral blood natural killer cells in children with autism’, Brain, behavior, and immunity, 23(1), pp. 124–133. Available at: 10.1016/J.BBI.2008.08.001.

17. Eyler, L.T., Pierce, K. and Courchesne, E. (2012) ‘A failure of left temporal cortex to specialize for language is an early emerging and fundamental property of autism’, Brain : a journal of neurology, 135(Pt 3), pp. 949–960. Available at: 10.1093/BRAIN/AWR364.

18. Feliciano, P., et al. (2019) ‘Exome sequencing of 457 autism families recruited online provides evidence for autism risk genes’, NPJ genomic medicine, 4(1). Available at: 10.1038/S41525-019-0093-8.

19. Gaugler, T., et al. (2014) ‘Most genetic risk for autism resides with common variation’, Nature genetics, 46(8), pp. 881–885. Available at: 10.1038/NG.3039.

20. Gazestani, V.H., et al. (2019) ‘A perturbed gene network containing PI3K-AKT, RAS-ERK and WNT-β-catenin pathways in leukocytes is linked to ASD genetics and symptom severity’, Nature neuroscience, 22(10), pp. 1624–1634. Available at: 10.1038/S41593-019-0489-X.

21. Grant, S.M. and Demorrow, S. (2020) ‘Bile Acid Signaling in Neurodegenerative and Neurological Disorders’, International Journal of Molecular Sciences 2020, Vol. 21, Page 5982, 21(17), p. 5982. Available at: 10.3390/IJMS21175982.

22. Gregg, J.P., et al. (2008) ‘Gene expression changes in children with autism’, Genomics, 91(1), pp. 22–29. Available at: 10.1016/J.YGENO.2007.09.003.

23. Hänzelmann, S., Castelo, R. and Guinney, J. (2013) ‘GSVA: gene set variation analysis for microarray and RNA-seq data’, BMC bioinformatics, 14. Available at: 10.1186/1471-2105-14-7.

24. He, Y., et al. (2019) ‘An integrated transcriptomic analysis of autism spectrum disorder’, Scientific reports, 9(1). Available at: 10.1038/S41598-019-48160-X.

25. Huang, H., et al. (2015) ‘Statistical Significance of Clustering using Soft Thresholding’, Journal of computational and graphical statistics : a joint publication of American Statistical Association, Institute of Mathematical Statistics, Interface Foundation of North America, 24(4), pp. 975–993. Available at: 10.1080/10618600.2014.948179.

26. Innis, S.M. (2007) ‘Dietary (n-3) Fatty Acids and Brain Development1’, The Journal of Nutrition, 137(4), pp. 855–859. Available at: 10.1093/JN/137.4.855.

27. Jia, W., et al. (2020) ‘Expert insights: The potential role of the gut microbiome-bile acid-brain axis in the development and progression of Alzheimer’s disease and hepatic encephalopathy’, Medicinal Research Reviews, 40(4), pp. 1496–1507. Available at: 10.1002/MED.21653.

28. Khokhlova, E. V., et al. (2020) ‘Features of DNA Repair in the Early Stages of Mammalian Embryonic Development’, Genes, 11(10), pp. 1–11. Available at: 10.3390/GENES11101138.

29. Kong, S.W., et al. (2012) ‘Characteristics and predictive value of blood transcriptome signature in males with autism spectrum disorders’, PloS one, 7(12). Available at: 10.1371/JOURNAL.PONE.0049475.

30. Kuwahara, A., et al. (2010a) ‘Wnt signaling and its downstream target N-myc regulate basal progenitors in the developing neocortex’, Development (Cambridge, England), 137(7), pp. 1035–1044. Available at: 10.1242/DEV.046417.

31. Kuwahara, A., et al. (2010b) ‘Wnt signaling and its downstream target N-myc regulate basal progenitors in the developing neocortex’, Development, 137(7), pp. 1035–1044. Available at: 10.1242/DEV.046417.

32. Lee, S.C., et al. (2019) ‘Solving for X: Evidence for sex-specific autism biomarkers across multiple transcriptomic studies’, American journal of medical genetics. Part B, Neuropsychiatric genetics : the official publication of the International Society of Psychiatric Genetics, 180(6), pp. 377–389. Available at: 10.1002/AJMG.B.32701.

33. Liberzon, A., et al. (2015) ‘The Molecular Signatures Database (MSigDB) hallmark gene set collection’, Cell systems, 1(6), pp. 417–425. Available at: 10.1016/J.CELS.2015.12.004.

34. Lombardo, M. V., et al. (2018) ‘Large-scale associations between the leukocyte transcriptome and BOLD responses to speech differ in autism early language outcome subtypes’, Nature neuroscience, 21(12), pp. 1680–1688. Available at: 10.1038/S41593-018-0281-3.

35. Lombardo, M. V. et al. (2021) ‘Atypical genomic cortical patterning in autism with poor early language outcome’, Science advances, 7(36). Available at: 10.1126/SCIADV.ABH1663.

36. Lord, C., et al. (2022) ‘The Lancet Commission on the future of care and clinical research in autism’, Lancet (London, England), 399(10321), pp. 271–334. Available at: 10.1016/S0140-6736(21)01541-5.

37. MahmoudianDehkordi, S., et al. (2019) ‘Altered bile acid profile associates with cognitive impairment in Alzheimer’s disease—An emerging role for gut microbiome’, Alzheimer’s & Dementia, 15(1), pp. 76–92. Available at: 10.1016/J.JALZ.2018.07.217.

38. Mainwaring, L.A., Bhatia, B. and Kenney, A.M. (2010) ‘Myc on my mind: a transcription factor family’s essential role in brain development’, Oncotarget, 1(2), p. 86. Available at: 10.18632/ONCOTARGET.113.

39. Marchetto, M.C., et al. (2017) ‘Altered proliferation and networks in neural cells derived from idiopathic autistic individuals’, Molecular psychiatry, 22(6), pp. 820–835. Available at: 10.1038/MP.2016.95.

40. McCarthy, M.M. (2008) ‘Estradiol and the developing brain’, Physiological Reviews, 88(1), pp. 91–124. Available at: 10.1152/PHYSREV.00010.2007/ASSET/IMAGES/LARGE/Z9J0040724530009.JPEG.

41. Min, Z., et al. (2019) ‘Asymmetrical methyltransferase PRMT3 regulates human mesenchymal stem cell osteogenesis via miR-3648’, Cell Death & Disease 2019 10:8, 10(8), pp. 1–17. Available at: 10.1038/s41419-019-1815-7.

42. Ní Ghrálaigh, F., et al. (2023a) ‘Brief Report: Evaluating the Diagnostic Yield of Commercial Gene Panels in Autism’, Journal of autism and developmental disorders, 53(1), pp. 484–488. Available at: 10.1007/S10803-021-05417-7.

43. Ní Ghrálaigh, F., et al. (2023b) ‘Brief Report: Evaluating the Diagnostic Yield of Commercial Gene Panels in Autism’, Journal of autism and developmental disorders, 53(1), pp. 484–488. Available at: 10.1007/S10803-021-05417-7.

44. Nutr, A. and Steiner, P. (2019) ‘Brain Fuel Utilization in the Developing Brain’, Annals of Nutrition and Metabolism, 75(Suppl. 1), pp. 8–18. Available at: 10.1159/000508054.

45. Ozsoy, S. (2020) ‘Heme as a Determinant of Brain Development’. Available at: https://ses.library.usyd.edu.au/handle/2123/21849 (Accessed: 18 November 2023).

46. Pertea, M., et al. (2015) ‘StringTie enables improved reconstruction of a transcriptome from RNA-seq reads’, Nature biotechnology, 33(3), pp. 290–295. Available at: 10.1038/NBT.3122.

47. Pierce, K., et al. (2019) ‘Evaluation of the Diagnostic Stability of the Early Autism Spectrum Disorder Phenotype in the General Population Starting at 12 Months’, JAMA Pediatrics, 173(6), pp. 578–587. Available at: 10.1001/JAMAPEDIATRICS.2019.0624.

48. Pramparo, T., Lombardo, M. V, et al. (2015) ‘Cell cycle networks link gene expression dysregulation, mutation, and brain maldevelopment in autistic toddlers’, Molecular systems biology, 11(12). Available at: 10.15252/MSB.20156108.

49. Pramparo, T., Pierce, K., et al. (2015) ‘Prediction of autism by translation and immune/inflammation coexpressed genes in toddlers from pediatric community practices’, JAMA psychiatry, 72(4), pp. 386–394. Available at: 10.1001/JAMAPSYCHIATRY.2014.3008.

50. Rashid, F., et al. (2017) ‘Induction of miR-3648 Upon ER Stress and Its Regulatory Role in Cell Proliferation’, International journal of molecular sciences, 18(7). Available at: 10.3390/IJMS18071375.

51. Redcay, E. and Courchesne, E. (2008) ‘Deviant functional magnetic resonance imaging patterns of brain activity to speech in 2-3-year-old children with autism spectrum disorder’, Biological psychiatry, 64(7), pp. 589–598. Available at: 10.1016/J.BIOPSYCH.2008.05.020.

52. Rumpf, S., Sanal, N. and Marzano, M. (2023) ‘Energy metabolic pathways in neuronal development and function’, Oxford open neuroscience, 2. Available at: 10.1093/OONS/KVAD004.

53. Satterstrom, F.K., et al. (2020) ‘Large-Scale Exome Sequencing Study Implicates Both Developmental and Functional Changes in the Neurobiology of Autism’, Cell, 180(3), pp. 568–584.e23. Available at: 10.1016/J.CELL.2019.12.036.

54. Stoner, R., et al. (2014) ‘Patches of disorganization in the neocortex of children with autism’, The New England journal of medicine, 370(13), pp. 1209–1219. Available at: 10.1056/NEJMOA1307491.

55. Taluja, V., et al. (2024) ‘Multimodality Integration of Neural Social Activation and Social and Language Scores Reveals Three Replicable Profound and Milder Autism Subtypes With Divergent Clinical Outcomes’, medRxiv, p. 2024.05.30.24308230. Available at: 10.1101/2024.05.30.24308230.

56. Turgeon, M.O., Perry, N.J.S. and Poulogiannis, G. (2018) ‘DNA Damage, Repair, and Cancer Metabolism’, Frontiers in oncology, 8(FEB). Available at: 10.3389/FONC.2018.00015.

57. Tylee, D.S., et al. (2017) ‘Blood transcriptomic comparison of individuals with and without autism spectrum disorder: A combined-samples mega-analysis’, American journal of medical genetics. Part B, Neuropsychiatric genetics : the official publication of the International Society of Psychiatric Genetics, 174(3), pp. 181–201. Available at: 10.1002/AJMG.B.32511.

58. Wang, B., et al. (2014) ‘Similarity network fusion for aggregating data types on a genomic scale’, Nature Methods 2014 11:3, 11(3), pp. 333–337. Available at: 10.1038/nmeth.2810.

59. Wen, T.H., et al. (2022) ‘Large scale validation of an early-age eye-tracking biomarker of an autism spectrum disorder subtype’, Scientific reports, 12(1). Available at: 10.1038/S41598-022-08102-6.

60. Weng, Z. Bin et al. (2022) ‘A Review of Bile Acid Metabolism and Signaling in Cognitive Dysfunction-Related Diseases’, Oxidative Medicine and Cellular Longevity, 2022. Available at: 10.1155/2022/4289383.

61. Xiao, Y., et al. (2022) ‘Neural responses to affective speech, including motherese, map onto clinical and social eye tracking profiles in toddlers with ASD’, Nature human behaviour, 6(3), pp. 443–454. Available at: 10.1038/S41562-021-01237-Y.

62. Xing, R. (2019) ‘miR-3648 Promotes Prostate Cancer Cell Proliferation by Inhibiting Adenomatous Polyposis Coli 2’, Journal of nanoscience and nanotechnology, 19(12), pp. 7526–7531. Available at: 10.1166/JNN.2019.16413.

63. Zhang, Q.G., et al. (2008) ‘Role of Dickkopf-1, an Antagonist of the Wnt/β-Catenin Signaling Pathway, in Estrogen-Induced Neuroprotection and Attenuation of Tau Phosphorylation’, Journal of Neuroscience, 28(34), pp. 8430–8441. Available at: 10.1523/JNEUROSCI.2752-08.2008.

64. Zhang, Y., Parmigiani, G. and Johnson, W.E. (2020) ‘ComBat-seq: batch effect adjustment for RNA-seq count data’, NAR genomics and bioinformatics, 2(3). Available at: 10.1093/NARGAB/LQAA078.

65. Zheng, X., et al. (2016a) ‘Metabolic reprogramming during neuronal differentiation from aerobic glycolysis to neuronal oxidative phosphorylation’, eLife, 5(JUN2016). Available at: 10.7554/ELIFE.13374.

66. Zheng, X., et al. (2016b) ‘Metabolic reprogramming during neuronal differentiation from aerobic glycolysis to neuronal oxidative phosphorylation’, eLife, 5(JUN2016). Available at: 10.7554/ELIFE.13374.

67. Zhou, X., et al. (2022) ‘Integrating de novo and inherited variants in 42,607 autism cases identifies mutations in new moderate-risk genes’, Nature genetics, 54(9), pp. 1305–1319. Available at: 10.1038/S41588-022-01148-2.

