## Supplementary Figures for "Beyond the Spectrum: Subtype-Specific Molecular Insights into Autism Spectrum Disorder Via Multimodal Data Integration"

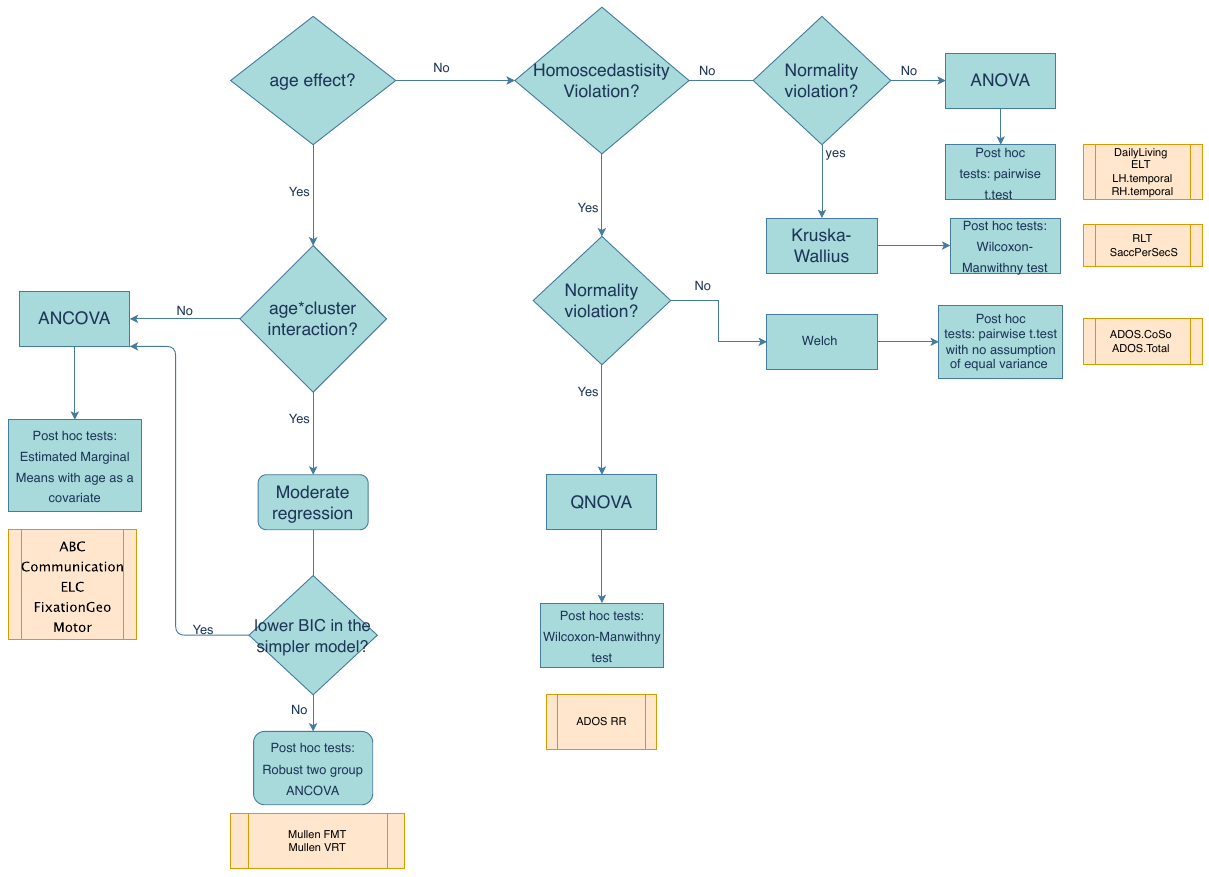

Figure S1: Selecting the appropriate statistical tests for group and pairwise comparisons. For age effects and interactions, ANCOVA is used when there is an age effect without interaction, while Bayesian Information Criteria (BIC) determines whether to include an age-cluster interaction term. If there is no age effect, we check normality using the Shapiro-Wilk test and homoscedasticity with Levene’s test. If assumptions are violated, the Kruskal-Wallis test or Welch test is used as alternatives. In cases of significant violations for both tests, Quantile-based ANOVA (QANOVA) is applied. Post-hoc analyses employ various tests based on assumption violations, including t-tests, Wilcoxon-Mann-Whitney tests, and robust ANCOVA methods.

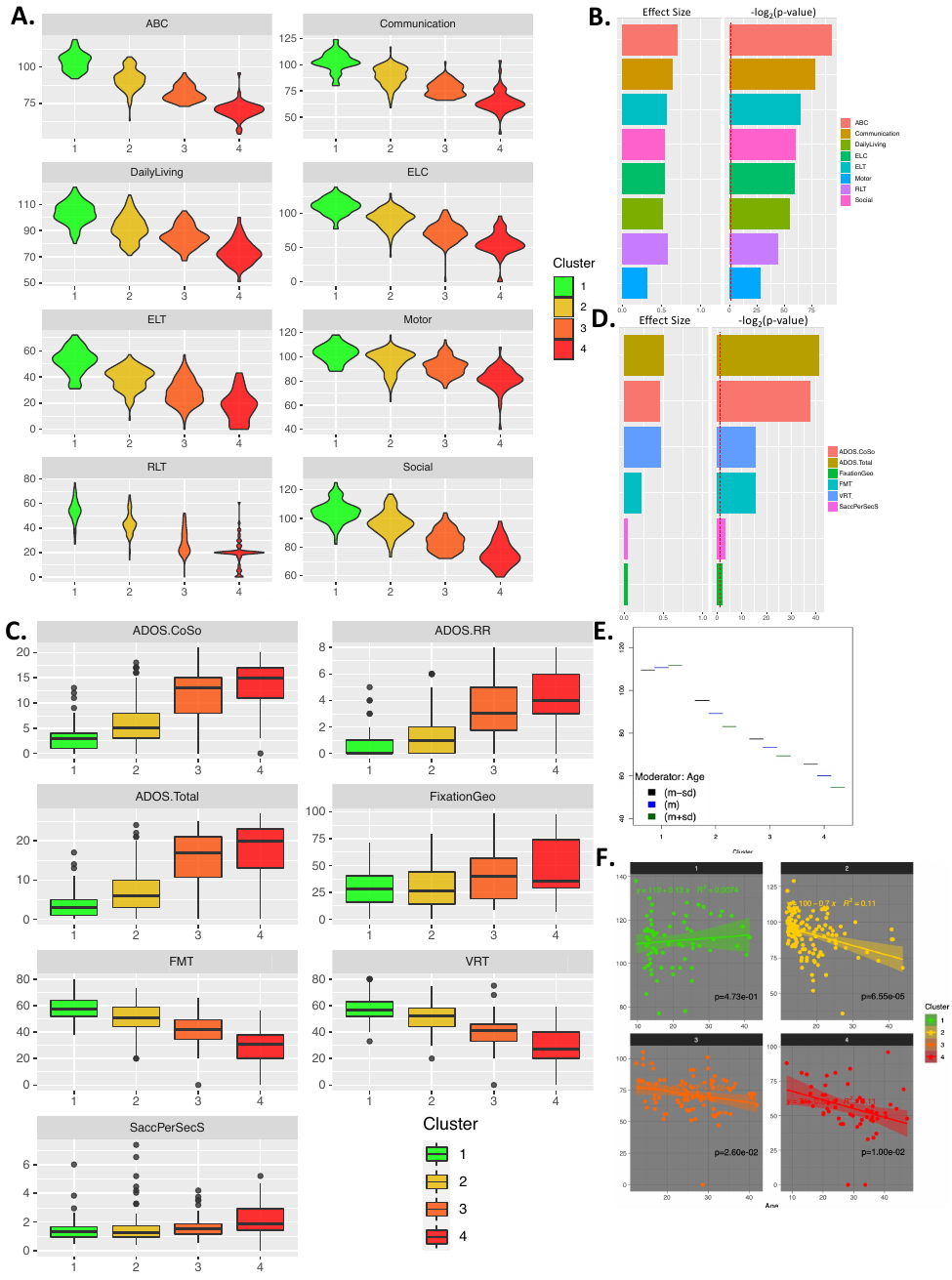

Figure S2

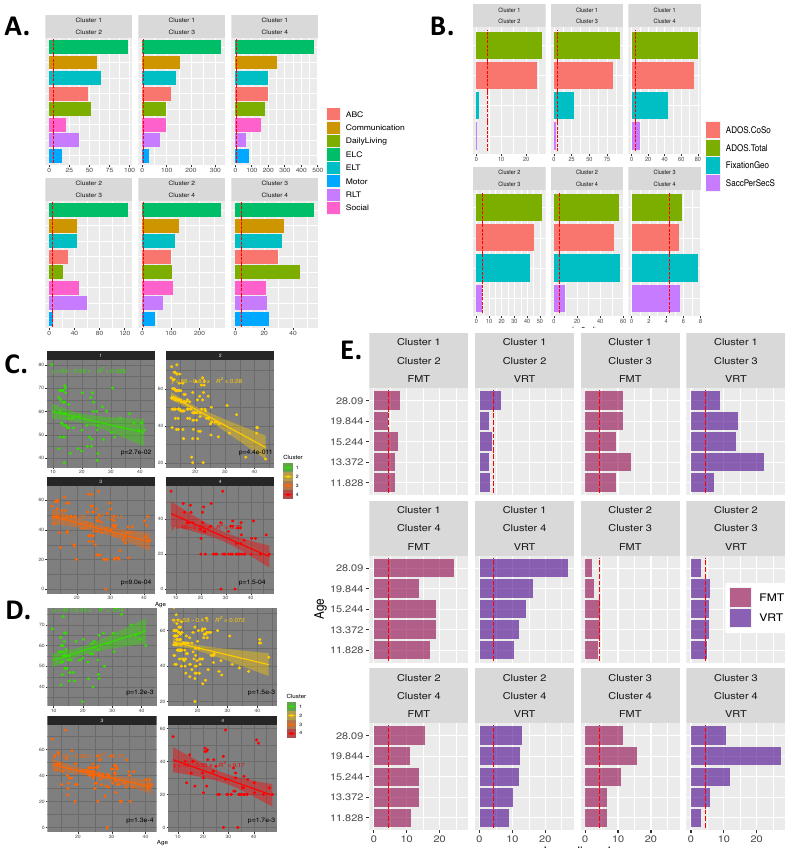

Figure S3

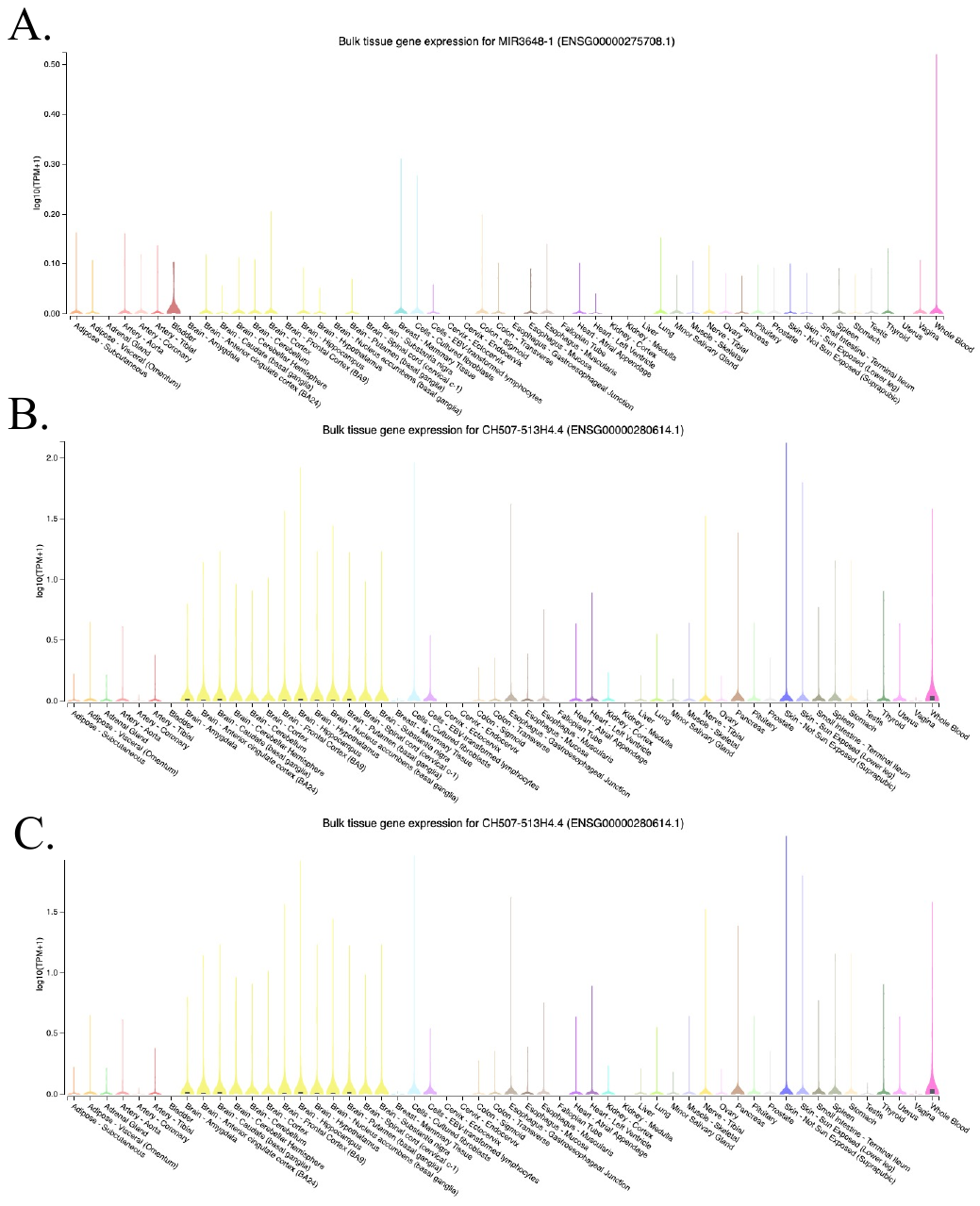

Figure S4

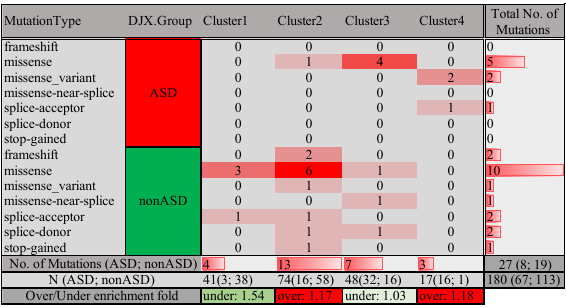

Figure S5

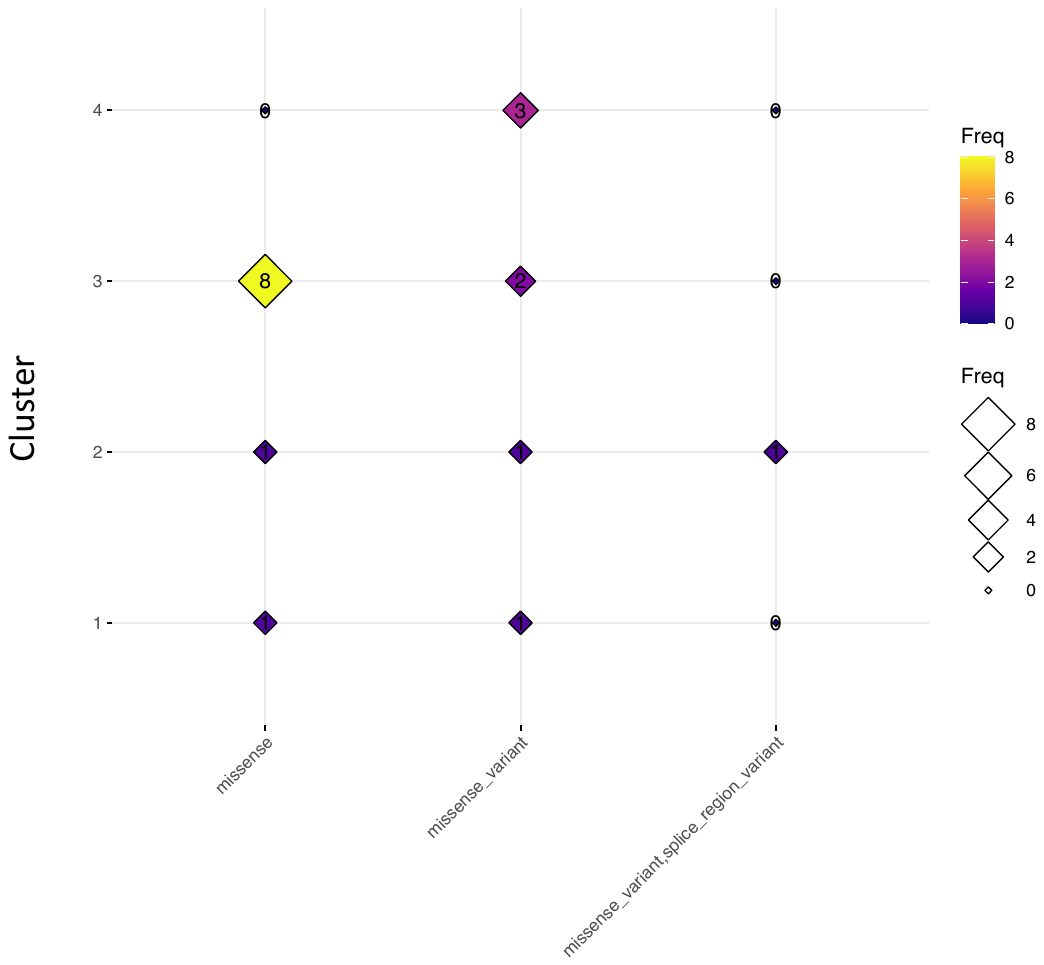

Figure S6
